# Cardiac SARS-CoV-2 infection is associated with distinct transcriptomic changes within the heart

**DOI:** 10.1101/2020.12.19.20248542

**Authors:** Diana Lindner, Hanna Bräuninger, Bastian Stoffers, Antonia Fitzek, Kira Meißner, Ganna Aleshcheva, Michaela Schweizer, Jessica Weimann, Björn Rotter, Svenja Warnke, Carolin Edler, Fabian Braun, Kevin Roedl, Katharina Scherschel, Felicitas Escher, Stefan Kluge, Tobias B. Huber, Benjamin Ondruschka, Heinz-Peter-Schultheiss, Paulus Kirchhof, Stefan Blankenberg, Klaus Püschel, Dirk Westermann

## Abstract

1

**Background:** Analyses in hospitalized patients and small autopsy series suggest that severe SARS-CoV-2 infection may affect the heart. We investigated heart tissue by in situ hybridization, immunohistochemistry and RNA sequencing in consecutive autopsy cases to quantify virus load and characterize cardiac involvement in COVID-19.

**Methods:** Left ventricular tissue from 95 deceased with diagnosed SARS-CoV-2 infection undergoing autopsy was analyzed and clinical data were collected. RNA was isolated to examine virus load of SARS-CoV-2 and its replication in the heart. A virus load >1000 copies per µg RNA was defined as relevant. Viral RNA and inflammatory cells were assessed using histology. RNA sequencing and gene ontology (GO) enrichment were performed in 10 cases with high cardiac virus load and 10 age-matched cases without cardiac infection.

**Results:** A relevant SARS-CoV-2 virus load was detected in 41 out of 95 deceased (43%). The median cardiac virus load was 7952 copies per µg RNA (IQR 2507, 32 005). In situ hybridization revealed SARS-CoV-2 RNA primarily in the interstitium or interstitial cells. Virus detection was not associated with increased inflammatory cells. Relevant cardiac infection was associated with increased expression of the entry factor *TMPRSS2*. Cardiac virus replication was found in 14/95 hearts (15%). Remarkably, cardiac virus replication was associated with shorter time between diagnosis and death. RNA sequencing revealed clear activation of immune response pathways to virus infection and destruction of cardiomyocytes. Hearts with high virus load showed activation of the GO term “extracellular exosomes”.

**Conclusion:** SARS-CoV-2 infection including virus replication and distinct transcriptomic alterations without signs of myocarditis demonstrate a cardiac involvement. In this autopsy series, cardiac replication of SARS-CoV-2 was associated with early death.

## 2 Introduction

Over 1.5 million people died in 2020 due to the global pandemic caused by SARS-CoV-2.^1^ Importantly, effective treatment options remain scarce due to limited knowledge of the exact pathological pathways involved in the disease. In addition to vaccinations, better therapies for patients with COVID-19 are needed. While repurposed antiviral drugs for patients hospitalized with COVID-19 did not reduce the overall mortality,^2^ immunosuppressant therapies such as dexamethasone and IgG infusions appear to improve outcomes in patients with COVID-19.^3, 4^ Therefore, understanding the immune response in organs that are key to survival may be essential for developing much needed treatment strategies.

While the virus is primarily targeting the respiratory system during the course of the disease other organs are infected as well.^5^ This results in thrombotic complications and direct injury of the kidneys and the heart.^6^ In the heart, inflammation can be detected by magnetic resonance imaging after COVID-19.^7^ Furthermore, cardiac dysfunction in patients with COVID-19 predicts a severe course of the disease and premature death,^8, 9^ additionally increased troponin serum levels are associated with death.^9, 10^ A previous pilot study from our group found SARS-CoV-2 viral RNA in 16/39 (41%) hearts of deceased who died with SARS-CoV-2 infection.^11^ It remains unclear whether this potentially dangerous cardiac involvement is the result of direct virus replication in the heart or secondary cardiac injury in the context of hypoxia, thrombosis, and multi-organ damage.^12^

To reveal cardiac infection with SARS-CoV-2 and associated pathological consequences, myocardial tissue was investigated directly in our studies.^11^ To address (i) the extent of virus infection and replication within the myocardial tissue, (ii) the virus localization in the heart and (iii) changes of gene expression associated with cardiac SARS-CoV-2 infection, we performed a systematic analysis of hearts obtained at autopsy from deceased who died with SARS-CoV-2 infection.

## 3 Material & Methods

### 3.1 Study cohort and tissue sampling

Consecutively, 95 deceased with proven SARS-CoV-2 infection were autopsied at the Institute of Legal Medicine at the University Medical Center Hamburg-Eppendorf in Germany between April and May, 2020. The SARS-CoV-2 infection was confirmed prior to death or post mortem by quantitative reverse transcription-polymerase chain reaction (RT-PCR) from pharyngeal swabs.^13^ Two tissue specimens from the free left ventricular wall were collected during autopsy and either snap frozen in liquid nitrogen or fixed in 10% neutral-buffered formalin for subsequent analyses.

This study was approved by the local ethics committee of the Hamburg Chamber of Physicians (PV7311). A brief report investigating the SARS-CoV-2 infection of the myocardial tissue for the first 39 cases already described a virus load of more than 1000 copies per µg RNA in 41% of the cases.^11^ This study was now extended to 95 cases and additional investigations. Furthermore, cases 1 to 39 were part of an autopsy study of first COVID-19 deaths in Hamburg, Germany, reporting causes of death and accompanying comorbidities.^14^

### 3.2 RNA isolation, reverse transcription and quantitative TaqMan PCR

Total RNA was isolated using QIAzol (Qiagen, Germany) and subsequently purified with the miRNeasy mini kit (Qiagen, Germany) as described earlier.^11, 15^ Reverse transcription of 1 µg RNA was carried out using the high-capacity complementary DNA (cDNA) kit (Thermo Fisher Scientific, USA) in the presence of random primers. Quantitative TaqMan PCR was performed using gene expression master mix (Thermo Fisher Scientific, USA). Specific gene expression assays are listed in Supplemental Table S1. Methods are described more detailed in the Supplemental Material.

### 3.3 Virus load copy number and replication

To determine the virus load in cardiac tissue, cDNA reversely transcribed with random primers was used. Quantitative TaqMan PCR was performed with primers and probe for the viral gene E as previously described.^16^ Copy number was calculated according to a purified and quantified PCR product and plotted as heat map.^11^

To detect virus replication, the intermediate minus RNA strand was transcribed into minus-strand-specific cDNA using the high-capacity cDNA kit either in the absence or in presence of 1 µM minus strand-specific primer. To avoid the detection of non-specific cDNA synthesis, the minus-strand specific primer E-Sarbeco-F was tagged by adding 14 nucleotides at the 5’ end (tagged-E-Sarbeco-F, 5’-TGCGATAATTGATACAGGTACGTTAATAGTTAATAGCGT-3’). This tagged minus-strand-specific cDNA was subsequently quantified by TaqMan PCR as described above using the primer shift-E-Sarbeco-F (5’-TGCGATAATTGATACAGGTACGTTAA-3’) instead of the non-modified E-Sarbeco-F primer. Tagging the primer for reverse transcription is known to avoid the detection of non-specific cDNA synthesis.^17, 18^ All primers and probes were purchased from Tib Molbiol, Germany. Virus replication was scored according to Ct-Values (0 – no replication, 1 – low replication, 2 – moderate replication, 3 – high replication) and plotted as heat map using Morpheus.^19^

### 3.4 Immunohistochemistry and immunofluorescence

Four micrometer-thick formalin-fixed paraffin embedded (FFPE) tissue sections were rehydrated and used for immunohistochemistry and immunofluorescence. Diagnostic immunohistological analyses were performed in the CAP-accredited laboratory IKDT (Institute for Cardiac Diagnostic and Therapy, Germany) as described in the Supplemental Methods and used for the characterization and quantification of inflammatory infiltrates. Myocardial inflammation was diagnosed by CD3^+^ T-lymphocytes/mm^2^, CD68^+^ macrophages/mm^2^, CD45R0^+^ memory T-cells, and CD8^+^ cytotoxic T-cells/mm^2^. Intramyocardial inflammation was assigned according to the European Society of Cardiology (ESC) statement by CD45R0^+^ cells > 14/mm^2^, CD3^+^ T-lymphocytes > 7/mm^2^, and in addition CD68^+^ cells > 14/mm^2^ and CD8^+^ cells > 1.8/mm^2^.^20^ To visualize increased inflammatory infiltrates, quantified immune cell numbers were plotted as x-fold to threshold of myocardial inflammation as heat map using Morpheus.^19^

Immunofluorescence was carried out according to established protocols and as described in the Supplemental Methods.^15, 21^

### 3.5 In situ hybridization – RNAscope

For detection of SARS-CoV-2 plus and minus strand, RNAscope in situ hybridization was performed on FFPE tissue sections of the left ventricle using RNAscope® 2.5 HD Reagent Kit-RED (#322350; Advanced Cell Diagnostics (ACD), USA). For fluorescent staining, RNAscope Multiplex Fluorescent v2 reagent kit (#323100, ACD, USA) was used. Tissue pretreatment, hybridization of target specific probe (listed in Supplemental Table S3), subsequent amplification steps and signal detection were performed according to the manufacturer’s instructions and described in detailed in the Supplemental Methods.

### 3.6 Massive analysis of cDNA Ends (MACE) as 3’mRNA Sequencing

For 3’mRNA sequencing analysis, 10 cases who exhibited a cardiac infection with more than 1000 copies per µg RNA were compared to 10 age-matched cases without cardiac infection selected from our study cohort. Massive Analysis of cDNA Ends (MACE) is a 3’mRNA sequencing method based on the analysis of Illumina reads derived from fragments that originate from 3’mRNA ends.^22^ The RNA samples were processed using the MACE-Kit v.2 according to the manufacturer’s protocol (GenXPro GmbH, Germany). Briefly, RNA was fragmented, polyadenylated mRNA was enriched and amplified by competitive PCR after poly-A specific reverse transcription and template-switch based second strand syntheses. Duplicate reads as determined by the implemented unique molecular identifiers (TrueQuant IDs) were removed from the raw dataset. Low quality sequence-bases were removed by the software cutadapt (https://github.com/marcelm/cutadapt/) and poly(A)-tails were clipped by an in-house Python-Script. The reads were mapped to the human genome (hg38) and transcripts were quantified by HTSeq.

Differential gene expression was calculated using DESeq2^23^ and plotted using Graph Pad Prism (GraphPad Software, USA). For the 19 most significant genes (FDR < 0.05), gene expression of all replicates (cases) was visualized as heat map. Therefore, normalized gene counts for each case were standardized to mean of the normalized gene counts of the control group (without cardiac infection) and plotted as log_2_-values. For data visualization R statistical software version 3.6.3 (R Foundation for Statistical Computing, Vienna, Austria) tool ComplexHeatmap was used.^24^

### 3.7 Gene Ontology Enrichment Analysis

Gene Ontology (GO) annotation data are based on ENSEMBL. GO enrichment analysis was performed using the topGO package (bioconductor). Enrichment of GO terms was calculated by the Fisher’s Exact Test based on differentially expressed transcripts (p-value < 0.05 and 2-fold up- or downregulated). The results of the GO enrichment analysis were visualized using the R package GOplot 1.0.2.^25^ The GO chord graph indicates associations between selected genes to selected GO terms and was generated utilizing GOChord. The left side of the circle displays selected genes together with their color-coded fold change (log_2_), whereas the right side of the circle shows selected GO terms. Genes and appertaining GO terms are connected by lines.

The GO circles were generated using GOCircle. The GO circle presents the differential gene expression for each annotated gene of the selected GO terms. In the outer circle, the fold change (log_2_) of all annotated genes within the GO term is displayed as scatter plot. While significantly upregulated genes are highlighted in red, significantly downregulated genes are highlighted in blue. In the inner circle, the significance of the GO term (-log_10_(p-value)) is indicated by the height of the bar chart, while its color displays the z-score. The z-score is calculated for each term by taking the number of upregulated genes, subtracting the number of downregulated genes, and dividing this number by the square root of the number of annotated genes. Whether the majority of genes in this term is up- or downregulated is displayed by the color gradient from red to blue, respectively.

### 3.8 Statistical analyses

Case characteristics were reported as medians and interquartile ranges in brackets for continuous variables. Binary variables are shown as counts with frequencies in brackets. The p-values were determined by the comparison to those cases with no cardiac infection. Mann-Whitney test was used for continuous variables and chi-squared test for binary variables.

To assess differences in outcome the primary endpoint of mortality was analyzed. Survival time of cases was estimated using the Kaplan-Meier method. A post-hoc pairwise comparison was conducted by the Log-rank test considering the Hochberg correction for multiple testing. To evaluate the association between comorbidities and cardiac infection, univariate logistic regression models were fitted. Forest plots display the respective odds ratio (OR) and 95% confidence interval (CI). The vertical line at an OR of one is the line of no effect. A clipped CI is indicated by arrows. A p-value of < 0.05 was considered statistically significant. All analyses were performed with R statistical software version 3.6.0 (R Foundation for Statistical Computing, Vienna, Austria).

## 4 Results

### 4.1 Cardiac infection of SARS-CoV-2

We investigated 95 deceased who died with a confirmed SARS-CoV-2 infection. As shown in Table 1, the median age was 84 years (IQR 76, 87) and 54% of them were female. The majority of cases died in hospitals (36% normal care and 26% intensive care) or nursing homes (27%, Supplemental Table S4). Cardiac tissue was collected during autopsy with a median post mortem interval of 4 days (IQR 3, 7). Virus load in cardiac tissue specimens was determined from isolated RNA by reverse transcription followed by quantitative PCR. As shown in Figure 2A, SARS-CoV-2 viral RNA in the heart was detected in 49 out of 95 cases (52%). In 46 cases, no SARS-CoV-2 was detected in cardiac tissue.

**Table 1:** Baseline characteristics for all 95 deceased and for subsequently defined groups of cases sorted by virus load or replication in cardiac tissue.

|  | All cases<br>(N=95) | Cases grouped by cardiac virus load or replication |  |  |  |  |
| --- | --- | --- | --- | --- | --- | --- |
|  |  | No cardiac infection<br>(N=46) | Cardiac infection<br>(> 1000 copies)<sup>†</sup><br>(N=41) | p-value* | Virus replication<br>(scored >1)<br>(N=14) | p-value* |
| **Female No. (%)** | 51 (53.7) | 21 (45.7) | 24 (58.5) | 0.32 | 7 (50.0) | 1.00 |
| **Age [years]** | 84.0 (76.0, 87.0) | 81.5 (74.2, 86.0) | 84.0 (78.0, 88.0) | 0.27 | 86.0 (82.5, 87.8) | 0.13 |
| **Post mortem interval [days]** | 4.0 (3.0, 7.0) | 4.0 (2.0, 6.0) | 4.0 (3.0, 8.0) | 0.38 | 5.0 (3.0, 9.8) | 0.17 |
| **Time, diagnosis until death [days]** | 9.0 (3.0, 18.0) | 12.5 (7.2, 20.2) | 8.0 (2.0, 10.0) | 0.082 | 2.0 (2.0, 9.0) | 0.017 |
| **Comorbidities** |  |  |  |  |  |  |
| HTN No. (%) | 44 (46.3) | 24 (52.2) | 17 (41.5) | 0.43 | 6 (42.9) | 0.76 |
| CAD No. (%) | 58 (61.1) | 31 (67.4) | 22 (53.7) | 0.28 | 7 (50.0) | 0.39 |
| HF No. (%) | 49 (51.6) | 23 (50.0) | 22 (53.7) | 0.90 | 8 (57.1) | 0.87 |
| **Sum of cardiac comorbidities ≥2** | 48 (50.5) | 26 (56.5) | 18 (43.9) | 0.34 | 8 (57.1) | 1.00 |
| DM No. (%) | 18 (18.9) | 11 (23.9) | 5 (12.2) | 0.26 | 4 (28.6) | 1.00 |
| Stroke No. (%) | 16 (16.8) | 7 (15.2) | 7 (17.1) | 1.00 | 5 (35.7) | 0.19 |
| COPD No. (%) | 49 (51.6) | 23 (50.0) | 24 (58.5) | 0.56 | 10 (71.4) | 0.27 |
| Cancer No. (%) | 29 (30.5) | 13 (28.3) | 14 (34.1) | 0.72 | 3 (21.4) | 0.87 |
| Renal failure No. (%) | 34 (35.8) | 16 (34.8) | 17 (41.5) | 0.67 | 8 (57.1) | 0.24 |
| **Sum of non-cardiac comorbidities ≥2** | 48 (50.5) | 22 (47.8) | 23 (56.1) | 0.58 | 11 (78.6) | 0.086 |
| **Sum of comorbidities ≥2** | 88 (92.6) | 42 (91.3) | 38 (92.7) | 1.00 | 14 (100) | 0.60 |
| **BMI [kg/m<sup>2</sup>]** | 24.4 (20.7, 28.7) | 24.4 (20.9, 28.4) | 23.2 (19.1, 28.7) | 0.58 | 25.0 (21.6, 26.8) | 0.85 |
| **Heart weight [g]** | 450.0 (361.2, 520.0) | 455.0 (383.8, 510.0) | 420.0 (333.8, 530.0) | 0.40 | 410.0 (330.0, 505.0) | 0.27 |

For continuous variables median (25<sup>th</sup> percentile, 75<sup>th</sup> percentile) are given. For binary variables absolute numbers and relative frequencies (%) are given.
\* The presented p-values were determined by the comparison to those cases without cardiac infection. Mann-Whitney test was used for continuous variables and chi-squared test for binary variables. The data availability for all variables is shown in the Supplemental Table S4.
<sup>†</sup> Since 1000 copies per $\mu\text{g}$ RNA was deemed as clinically relevant, 8 out of 95 cases who not exceed 1000 copies per $\mu\text{g}$ RNA were excluded from the group with cardiac infection.
Abbreviations: BMI, body mass index; CAD, coronary artery disease; COPD, chronic obstructive pulmonary disease; DM, diabetes mellitus; HF, heart failure; HTN, hypertension;
No, numbers.

**Figure 1.**
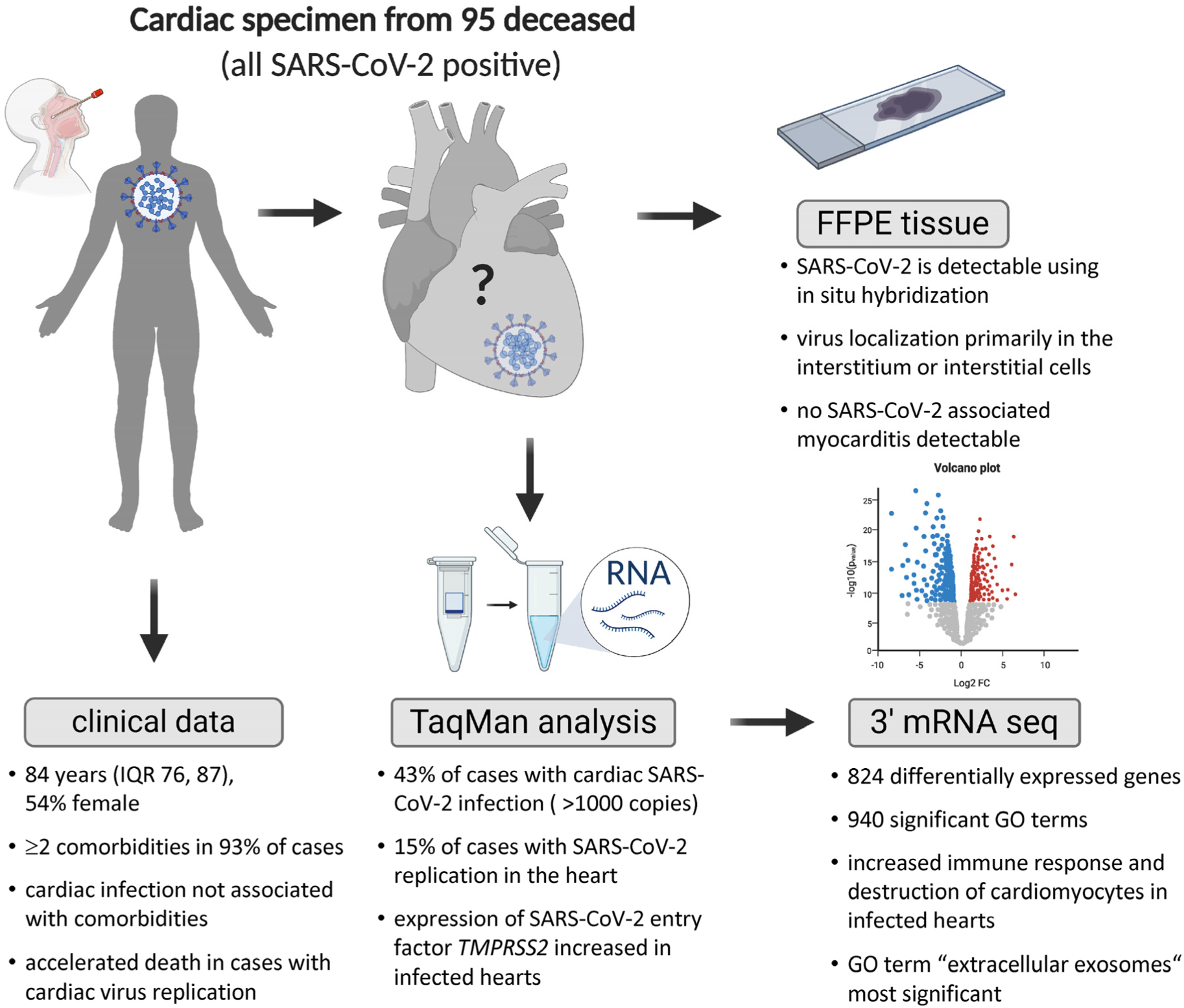
Central Figure: Study cohort and study design. Two cardiac tissue specimens per case underwent molecular and histological analyses. Virus load and its replication was determined from isolated RNA. Based on the presence of viral RNA and virus replication in the myocardium, cases were grouped and compared. While comorbidities were not associated with virus presence or replication in the heart, more rapid death appeared in those cases with virus replication in the heart. On formalin-fixed paraffin embedded (FFPE) tissue slices, SARS-CoV-2 RNA was visualized primarily in the interstitium or interstitial cells. This was not associated with an influx of inflammatory cells. For 3’mRNA sequencing, 10 cases with high virus load in cardiac tissue were compared to 10 age-matched cases without cardiac infection. Transcriptional profiling allowing for gene ontology (GO) enrichment analyses revealed clear immune response to virus infection in the myocardium.

**Figure 2.**
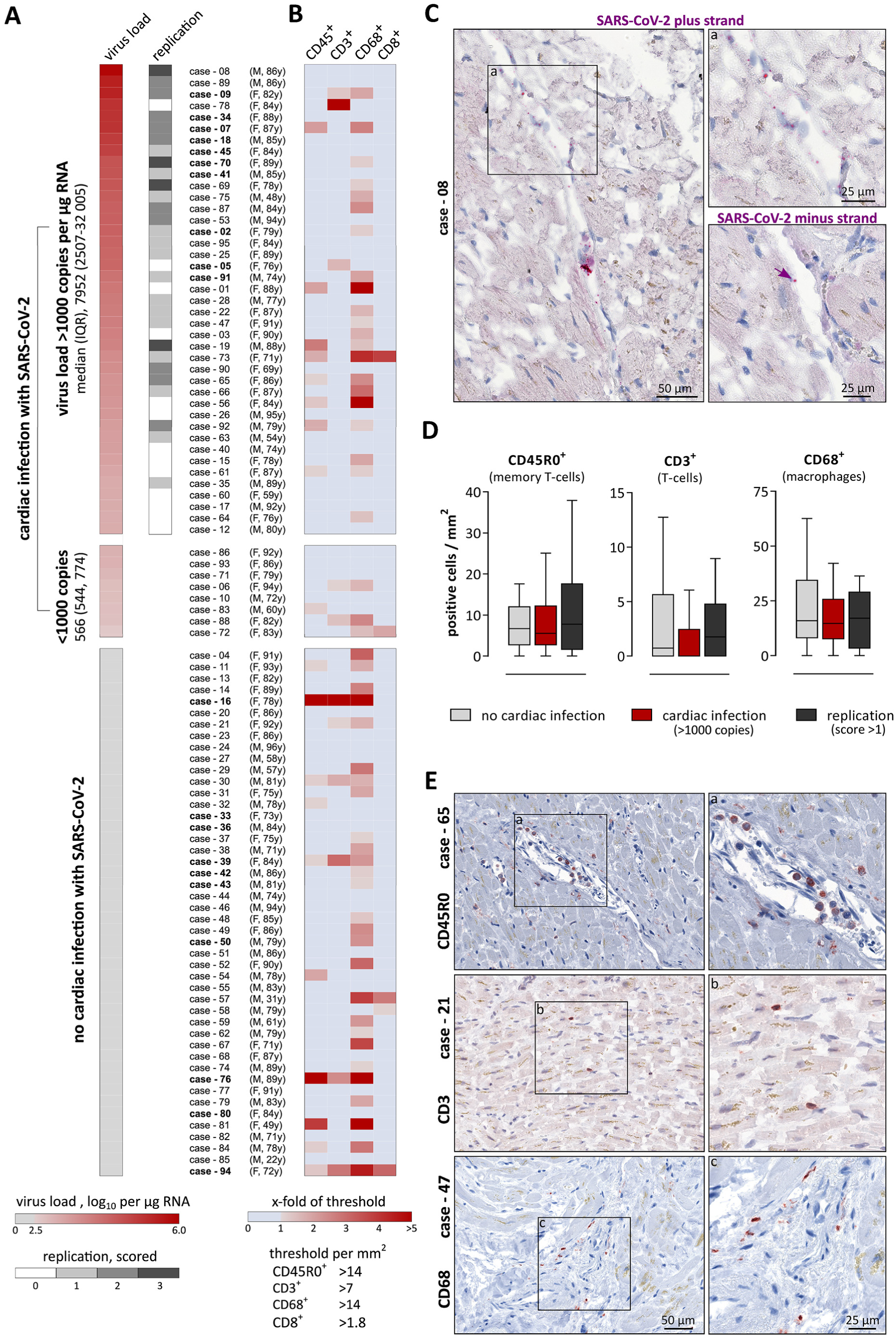
Presence and replication of SARS-CoV-2 in cardiac tissue is not associated with elevated number of inflammatory cells in deceased died with SARS-CoV-2 infection. A: Presence of SARS-CoV-2 RNA was examined in the cardiac tissue of 95 SARS-CoV-2 positive deceased. Autopsy cases are sorted by determined cardiac virus load (shown in red). Virus replication is shown in grey. For consecutive case-numbers, age and sex are presented in brackets. Virus load in 1 µg RNA was quantified by reverse transcription followed by quantitative polymerase chain reaction (PCR). A copy number of more than 1000 copies per µg RNA was deemed as clinically relevant and detected in 41 cases, which were further examined for virus replication. Virus replication was determined using a tagged minus-strand specific primer for reverse transcription followed by quantitative PCR of the tagged cDNA. Replication was scored according to the Ct-values (0-no replication / 1-weak / 2-moderate / 3-strong). Note: cases further analyzed using 3’mRNA-sequencing are highlighted in bold. B: Immunohistochemistry staining was performed on formalin- fixed paraffin embedded (FFPE) tissue specimens of the myocardium. For each case individually, cells positive for the respective immune cell marker were quantified and plotted as x-fold to threshold numbers described for diagnosing myocarditis (Dallas criteria). Red indicates increased immune cell number. C: In situ hybridization was used to visualize SARS-CoV-2 RNA on FFPE tissue specimens. Hybridizing probes are specific either for the plus strand representing the virus genome or the minus strand representing the intermediate strand for replication. Representative images of case - 08 are displayed. D: The number of positively stained immune cells per mm^2^ is displayed. Cases without cardiac infections are depicted in light grey, whereas cases with cardiac infections higher than 1000 copies per µg RNA are depicted in red. Cases with cardiac infection were constricted to those with virus replication scores higher than 1 and are depicted in dark grey. Both groups with cardiac infections were separately compared to the group without cardiac infection. Data are depicted as Tukey-style box plots with median and interquartile range without outliers. E: Representative staining of immune cells in cardiac tissue are depicted. On the upper panel, the CD45R0 staining for case - 65, quantified as 15.14 cells per mm^2^, is shown. The middle panel depicts the CD3 staining for case - 21 (7.37 cells / mm^2^). For case - 47, the CD68 staining is shown on the lower panel (14.72 cells / mm^2^). Magnifications are displayed for each case on the right side. (F, female; IQR, interquartile range; M, male; y, years)

Since 1000 copies per µg RNA was deemed as clinically significant,^11^ 41 out of 95 cases (43%) who exceed 1000 copies per µg RNA were included in further analyses. This group revealed a median virus load of 7952 copies per µg RNA (IQR 2507, 32 005). In 28 out of these 41 cases with cardiac infection, virus replication was detectable, 14 with weak, 10 with moderate and 4 with strong virus replication. Additionally, the virus was visualized in tissue sections utilizing in situ hybridization. As shown in Figure 2C, SARS-CoV-2 virus genome (plus strand) was detected frequently, whereas SARS-CoV-2 intermediate strand for replication (minus strand) was rarely detectable in the consecutive section. In this representative case, the virus genome was apparently detectable in non-cardiomyocyte structures. To investigate the localization of virus particles, we performed fluorescent co-staining of SARS-CoV-2 plus stand (red) and wheat germ agglutinin (WGA, cyan) as marker for cell membranes and fibrotic tissue (Figure 3). Negative and positive controls for chromogenic and fluorescent in situ hybridization are shown in the Supplemental Figure S1 and S2, respectively. In the majority of cases, virus particles are co-localized with WGA indicating a virus localization in the interstitium or in interstitial cells. Localization within cardiomyocytes as depicted in Figure 3g is rare.

**Figure 3:**
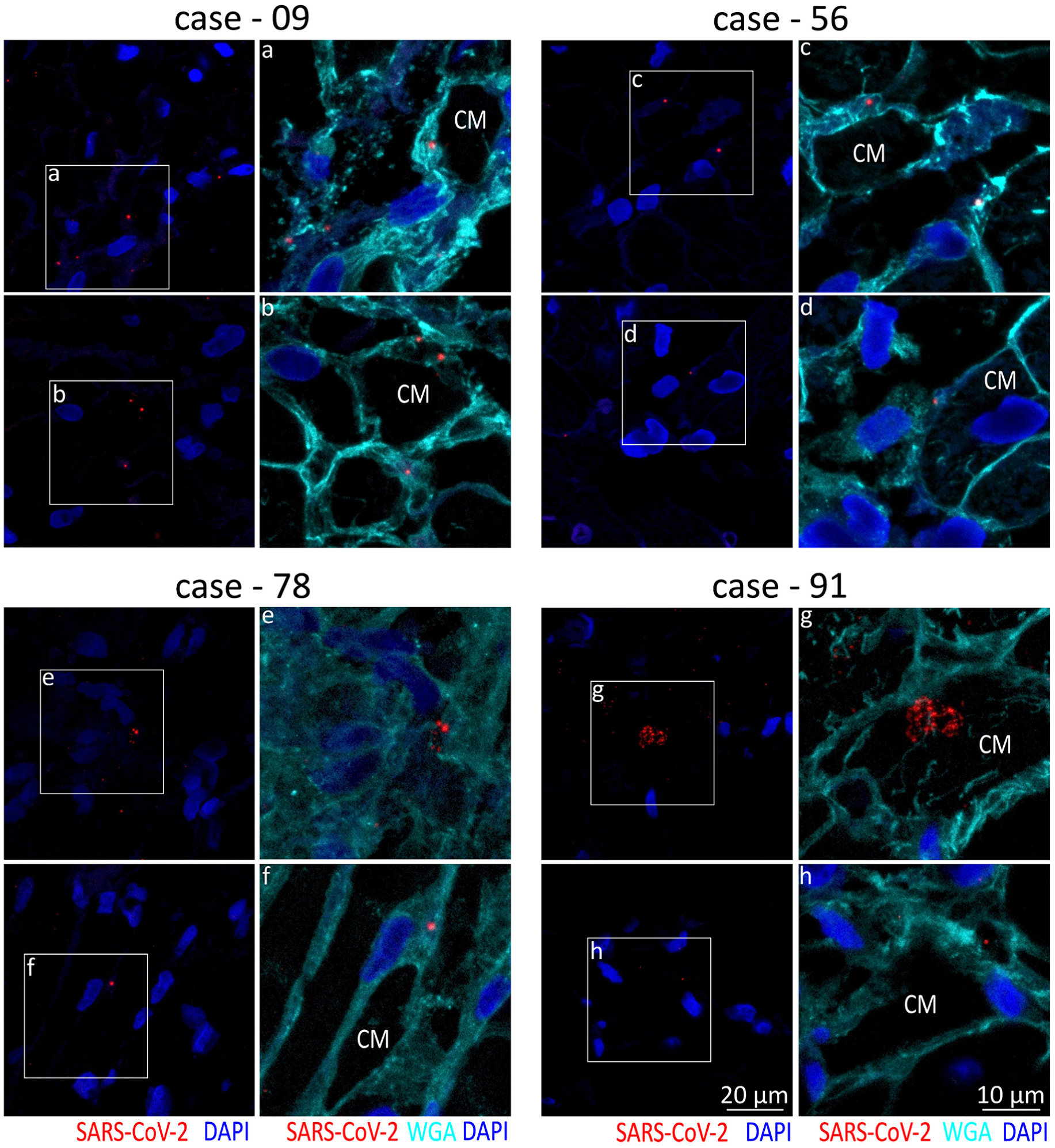
In situ hybridization of SARS-CoV-2 RNA in cardiac tissue revealed localization in the interstitium or interstitial cells. Two representative fluorescence staining on formalin-fixed paraffin embedded cardiac tissue from four different cases are shown. All four cases exhibited a virus load higher than 1000 copies per µg RNA. Red signal indicates SARS-CoV-2 plus strand, cell nuclei are stained in blue. White boxes indicate zoomed in areas displayed on the right. The zoomed in image additionally displays co-staining with wheat germ agglutinin (WGA, cyan), which stains the interstitium or interstitial cells and surrounds the large cardiomyocytes (CM). In case - 09, - 56 and - 78 the virus particles are co-localized with WGA close to cardiomyocytes, which represent the finding in the majority of cases. Localization of virus particles within cardiomyocytes as observed in case - 91 is rare.

Next, histological signs of immune cell infiltrates were determined by immunohistochemistry. Therefore, CD45R0^+^ memory T-cells, CD3^+^ T-cells, CD68^+^ macrophages and CD8^+^ cytotoxic T-cells were quantified and plotted for each case individually in Figure 2B. Individuals with elevated numbers of infiltrated cells were not differentially distributed in cases with or without cardiac infection. In addition, the number of positive cells were compared between those cases without cardiac infection and those with cardiac infection higher than 1000 copies per µg RNA or to those with clearly detectable virus replication (Figure 2D). No significant differences were determined for CD45R0, CD3 and CD68 between both groups, while CD8 was only detectable in very few cases. Representative staining are shown for three different cases representing increased infiltrated cells of distinct subpopulations in Figure 2E.

### 4.2 SARS-CoV-2 infection of the heart is not associated with comorbidities

According to virus load and virus replication of SARS-CoV-2 in cardiac tissue, we defined the group without cardiac infection and the group with cardiac infection higher than 1000 copies which was further constricted to cases with virus replication scores higher than 1. Two comparisons were performed: (i) 46 cases without cardiac infection to 41 cases with cardiac infection higher than 1000 copies per µg RNA and (ii) 46 cases without cardiac infection to 14 cases with virus replication scores higher than 1. As shown in Table 1, both comparisons revealed no significant differences in age, sex and post mortem intervals indicating matched groups with respect to typical confounders in post mortem setting. However, time between diagnosis and death, representing the interval between the first positive respiratory swab for SARS-CoV-2 and date of death, was significantly smaller in cases with virus replication in the heart compared to cases without cardiac infection (p = 0.017). Accordingly, time between diagnosis and death was analyzed utilizing Kaplan-Meier Curves shown in Figure 4A and revealed significant differences as well for comparison of replication positive cases with samples without cardiac infection (p = 0.012). Virus replication in the heart was associated with a significantly shorter survival time.

**Figure 4:**
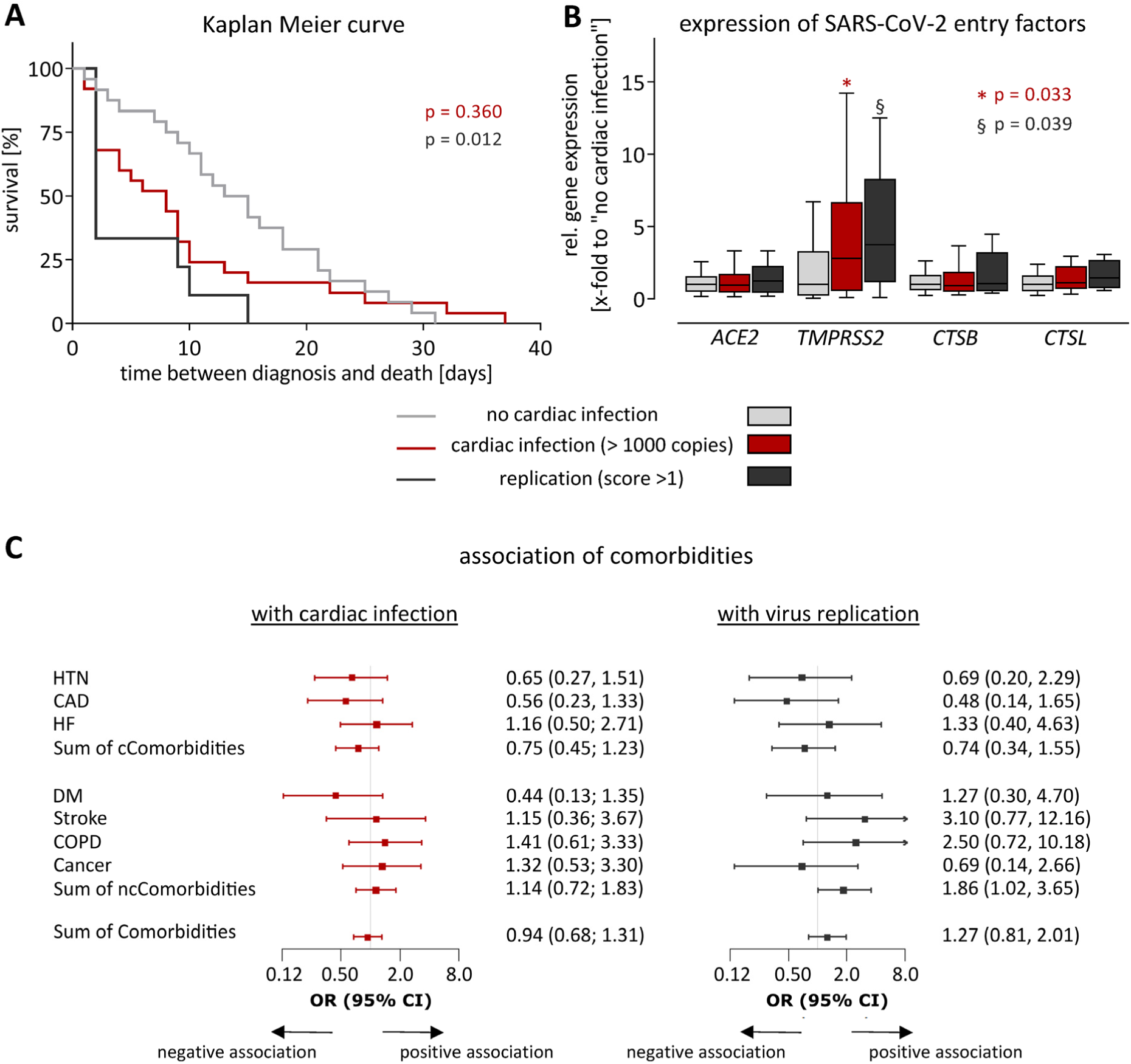
Cardiac infection with SARS-CoV-2 is associated with accelerated death but not with comorbidities. Cases without cardiac infection are depicted in light grey, whereas cases with cardiac infection higher than 1000 copies per µg RNA are depicted in red. Cases with cardiac infection were constricted to those with virus replication scores higher than 1 and are depicted in dark grey. Both groups with cardiac infection were separately compared to the group without cardiac infection. A: Kaplan-Meier curves show time between diagnosis and death for cases grouped by cardiac infection and virus replication. Time between diagnosis and death is given by the interval between the first positive respiratory swab for SARS-CoV-2 and date of death. The compared groups were defined using information that was unknown before death. Both groups with cardiac infection were compared to the group without cardiac infection utilizing log-rank test. To compare multiple survival curves, p-values were adjusted by the Hochberg correction. B: Expression of four SARS-CoV-2 entry genes was determined using TaqMan analysis. Gene expression is plotted normalized to the median expression of the group with no cardiac infection. Data are depicted as Tukey-style box plots with median and interquartile range without outliers. Significant differences were determined using Mann-Whitney Test. A p-value of < 0.05 was considered statistically significant. C: Comorbidities and cardiac infection or replication were fitted using univariate logistic regression models. Odds ratios (OR) and 95% confidence intervals (CI) were displayed as forest plots. The vertical line at an OR of one is the line of no effect. Due to graphical issues the CI is clipped indicated by arrows. A p-value of < 0.05 was considered statistically significant, but not reached for this analysis. Abbreviations: CAD, coronary artery disease; cCormobidites, cardiac Comorbidities; COPD, chronic obstructive pulmonary disease; DM, diabetes mellitus; HF, heart failure; HTN, hypertension; ncCormobidities, non cardiac comorbidities.

Next, we examined whether cardiac infection or virus replication is associated with expression levels of entry factors described for SARS-CoV-2. Therefore, gene expression of angiotensin-converting enzyme-2 (*ACE2*), transmembrane serine protease-2 (*TMPRSS2*), cathepsin-B (*CTSB*) and cathepsin-L (*CTSL*) was compared. As depicted in Figure 4B, gene expression of *TMPRSS2* was significantly increased in groups with cardiac infection and replication, whereas gene expression of *ACE2, CTSB* and *CTSL* remained nearly unchanged.

During autopsy, comorbidities of each case were diagnosed. To analyze whether comorbidities promote cardiac infection, odds ratios for the cardiac comorbidities hypertension (HTN), coronary artery disease (CAD), heart failure (HF) or the sum of cardiac comorbidities revealed no association (Figure 4C) between comorbidities and cardiac infection or replication. Additionally, other non cardiac comorbidities such as diabetes mellitus (DM), stroke, chronic obstructive pulmonary disease (COPD), cancer or the sum of non cardiac or all comorbidities are not associated with cardiac infection.

### 4.3 Identification and gene ontology analyses of top differentially expressed genes using 3’mRNA sequencing

For 3’mRNA sequencing, 10 cases with high virus load in cardiac tissue were selected and compared to 10 age-matched SARS-CoV-2 positive cases without cardiac infection (Supplemental Table S4). As shown in Figure 5A, 23 335 genes were annotated whereof 824 genes were identified as significantly different. Out of these 824 genes, 394 genes were significantly upregulated (red) and 430 genes significantly downregulated (blue) in cardiac tissue with high virus load of SARS-CoV-2. Next, we aimed to compare regulated genes with protein expression. Olink Proteomics disseminate a publicly available data set analyzing over 1400 plasma proteins in a COVID-19 cohort including symptomatic controls.^26^ Out of the 824 differentially expressed genes in our cohort, 73 corresponding plasma proteins were measured in the Olink data set. Out of these 73 targets, 48 were up- and 25 were downregulated on gene expression level in infected myocardial tissue. Comparing plasma protein levels of refined groups with and without COVID-19, overlaps between gene expression and plasma levels were identified. Out of the 48 myocardial upregulated genes, 16 were confirmed to be elevated on plasma protein level in COVID-19 patients. (For more detail see Supplemental Material and Supplemental Figure S3.)

**Figure 5:**
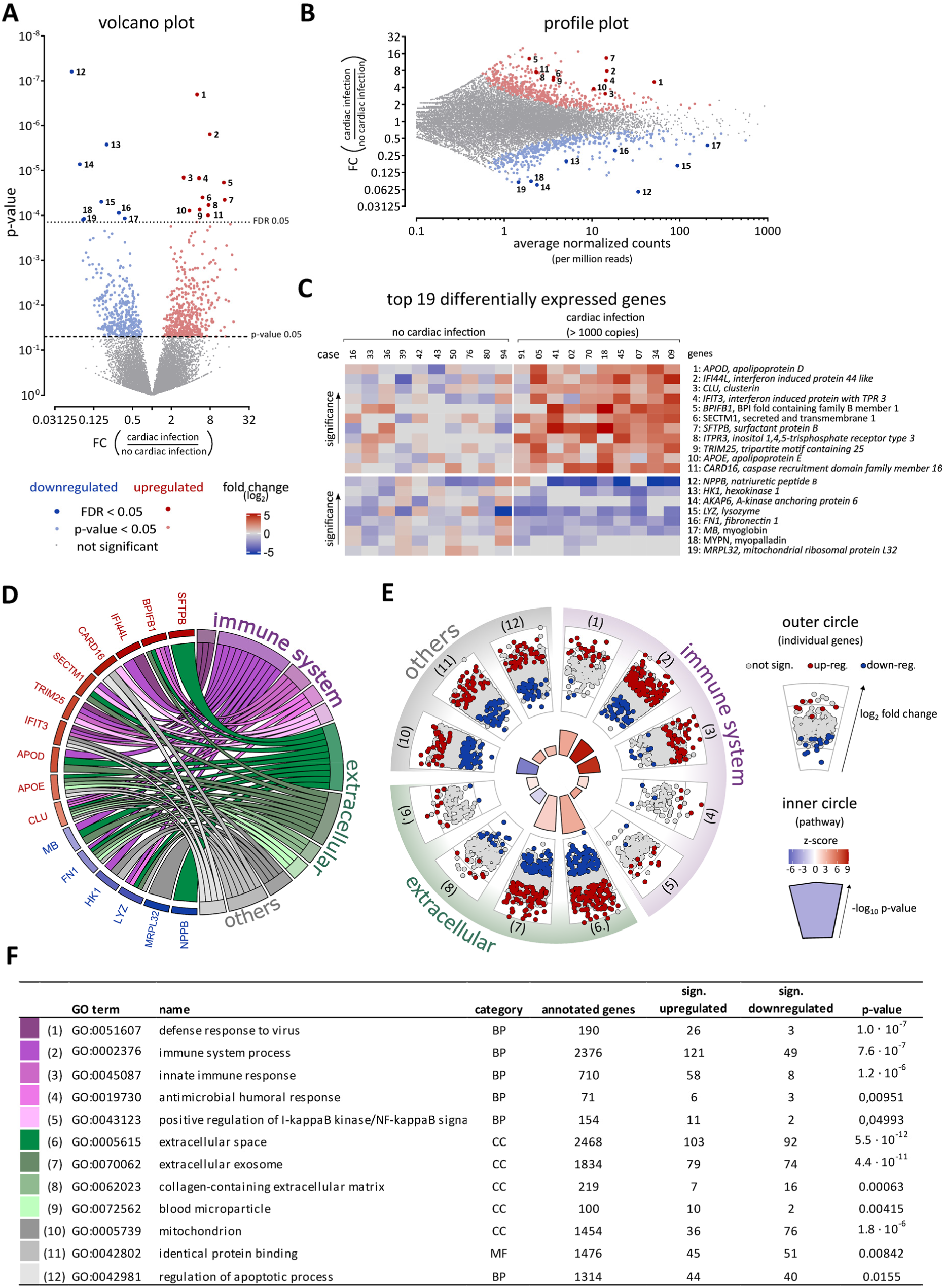
Identification and gene ontology analyses of the top 19 differentially expressed genes. Gene expression measured by 3’mRNA sequencing of 10 cases with cardiac SARS-CoV-2 infection was compared to 10 cases without cardiac infection. Downregulated genes in SARS-CoV-2 infected cardiac tissue are depicted in blue, while red highlights upregulated genes. A: Fold change and p-value are displayed in the volcano plot to visualize significant differences in gene expression. Further restricted by a false discovery rate (FDR) < 0.05, 19 top differentially expressed genes were identified (11 upregulated and 8 downregulated genes). Gene symbols and full names are displayed in C according to here depicted numbers. B: In the profile plot, fold change and average of normalized counts per 1 million reads are displayed. Red and blue colors highlight significantly regulated genes with p-value < 0.05 (small dots) or FDR < 0.05 (large dots). Gene symbols and full names are displayed in C according to here depicted numbers. C: The top 19 differentially expressed genes are displayed for each case separately. Gene symbols and full names are given. Gene expression is normalized to the mean of the group without cardiac infection and plotted as fold change (log_2_). D: In the gene ontology (GO) chord graph, the left side of the circle displays the top 19 genes together with their color-coded fold change. The right side of the circle shows the GO terms, to which the genes are annotated. Only significant GO terms including at least 3 of the top 19 genes are presented, large GO terms including more than 3000 annotated gens are not displayed. Genes and appertaining GO terms are connected by lines. Color codes for a specific GO term and its function. E: The GO circle presents the differential gene expression for each annotated gene of the GO terms identified by the GO chord in panel D. In the outer circle, the fold change of all annotated genes within the GO term is displayed. While significantly upregulated genes are highlighted in red, significantly downregulated genes are highlighted in blue. In the inner circle, the significance (-log_10_(p-value)) is indicated by the height of the bar chart, while its color displays the z-score. Whether the majority of genes in this term is up- or downregulated is displayed by the color gradient from red to blue, respectively. All significantly regulated genes are listed in the Supplemental Table S5. F: The list presents details about the GO terms shown in D&E. The color code in the first column indicates the GO term in panel D, while the number in the second column indicates the GO term in panel E.

The 824 differentially expressed genes were further restricted by the false discovery rate (FDR) < 0.05 resulting in top 19 differentially expressed genes, 11 upregulated and 8 downregulated. The most significantly upregulated gene is *APOD* (No. 1) while the most significantly downregulated gene is *NPPB* (No. 12). As depicted in the profile plot, both are abundantly expressed in the cardiac tissue, whereas the upregulated genes *BPIFB1* (5), *ITPR3* (8), *CARD16* (11) and the downregulated genes *MRPL32* (19), *MYPN* (18), *AKAP6* (14) are rarely expressed (Figure 5B). The data and names for all top 19 differentially expressed genes are plotted for each case separately as heat map in Figure 5C.

Next, GO enrichment analyses were performed to identify significant GO terms in infected myocardiums. We selected all GO terms which include at least 3 of the top 19 genes. Afterwards, non-significant GO terms as well as large GO terms with more than 3000 annotated genes were excluded. Subsequently, we identified 12 significantly regulated GO terms which are depicted in the GO chord graph in Figure 5D and listed in 5F. 9 out of these 12 GO terms were linked to the immune system (5 terms, displayed in purple) or to extracellular regions (4 terms, displayed in green), whereas the remaining 3 GO terms were displayed in grey. Furthermore, 15 out of the top 19 genes are annotated in GO terms linked to the immune system.

The detailed results for the 12 GO terms displayed in the GO chord are plotted in Figure 5E and F. The majority of genes in the GO terms linked to the immune system (1-5) were upregulated, indicated by red z-scores. Remarkably, “defense response to virus” (1) is the most significant GO term in this group and emphasizes the SARS-CoV-2 infection of the cardiac tissue. In the extracellular group, the GO terms extracellular space (6) and extracellular exosome (7) reach the highest significance. In the highly significantly regulated GO term mitochondrion (10), most genes are downregulated, indicated by a blue z-score.

### 4.4 Untargeted and targeted selection of GO terms

GO term analyses of the 3’mRNA sequencing data revealed 940 significantly regulated GO terms within all three GO categories, 98 in molecular function (MF), 159 in cellular component (CC) and 683 in biological process (BP). To narrow these GO term analyses we performed an untargeted and a targeted selection.

First, for each of the three categories BP, CC and MF ancestor charts of the 15 most significant GO terms were created. Since the significance of parental and child terms is often based on the same set of genes, we selected most descendent child terms with at least 40 annotated genes for each category. Overall, 11 child terms were obtained and displayed in Figure 6A. The GO terms were linked to general topics such as (i) response to virus, (ii) cardiomyocyte structure, and (iii) respiratory chain. GO terms (1-3) linked to (i) response to virus almost exclusively include significant upregulated genes which is further emphasized by the red z-scores. In contrast, GO terms (9-11) linked to (ii) cardiomyocyte structure and GO terms (6-8) linked to (iii) respiratory chain contain primarily significant downregulated genes which implies the loss of cardiomyocyte structure and mitochondrial function in cardiac tissue infected with SARS-CoV-2.

**Figure 6:**
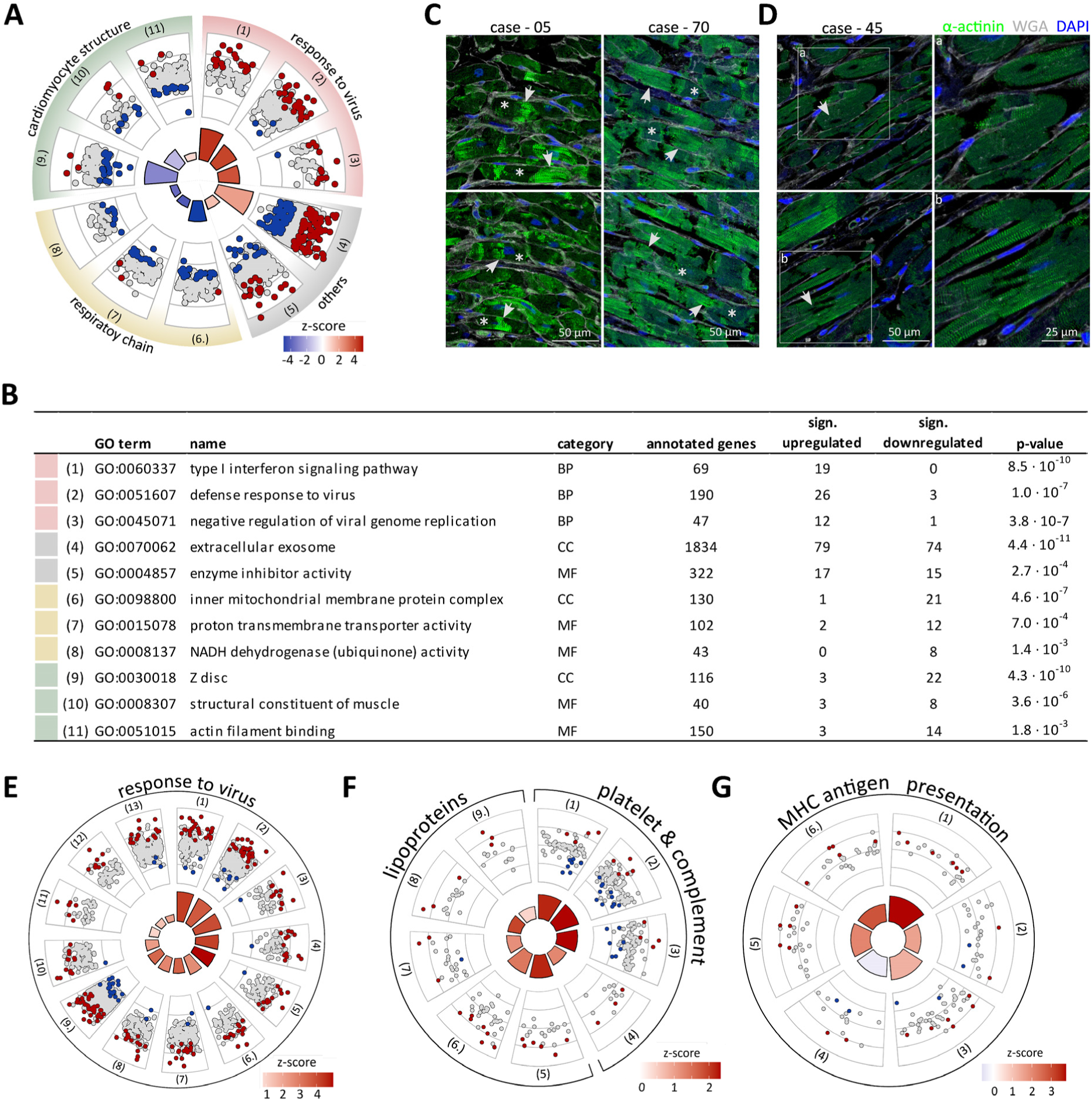
Gene Ontology (GO) enrichment analyses indicate response to virus and impaired cardiomyocyte structure in the SARS-CoV-2 infected cardiac tissue. A: GO circle of the most significant GO terms. Therefore, we selected the most descendent child terms in the ancestor chart displaying the 15 most significant GO terms. The corresponding ancestor charts for the three categories are shown in the Supplemental Figures S4-S6. The GO circle presents the results for each GO term separately. In the outer circle, the fold change of all annotated genes within the GO term is displayed. While significantly upregulated genes are highlighted in red, significantly downregulated genes are highlighted in blue. In the inner circle, the significance (-log_10_(p-value)) is indicated by the height of the bar chart, while its color displays the z-score. Whether the majority of genes in this term is up- or downregulated is displayed by the color gradient from red to blue, respectively. All significantly regulated genes are listed in the Supplemental Table S6. B: The list presents details about the GO terms shown in panel A. Color-codes in the first column indicate the general topic to which the term is linked, while the number in the second column indicates the numbered GO term in panel A. C: Two representative α-actinin staining (green) of cardiac tissue section from case - 05 and case - 70 (both exhibiting high virus load of SARS-CoV-2 in the heart) are shown. The normal sarcomeric structure of cardiomyocytes is indicated by arrows, whereas inconsistent actinin staining as a sign of sarcomeric disarray is indicated by asterisks. Nuclei counterstained with DAPI are shown in blue, extracellular matrix stained with WGA are shown in white. D: Two representative images of cardiac tissue from case - 45 who exhibited high virus load in the heart. White boxes indicted zoomed in areas of the image on the right. Some cardiomyocytes lack DAPI staining for the nuclei. White arrows denote the putative region of nuclear localization. E-G: According to the current literature, the list of 940 significant GO terms was searched for specific keywords. E: 13 GO terms were identified within the 940 significant GO terms searched for keywords “virus” and “viral”. Detailed results and significantly regulated genes are listed in the Supplemental Table S7. Red z-scores indicate upregulated gene expression in SARS-CoV-2 infected cardiac tissue. F: 4 significant GO terms (1-4) were identified for keywords “platelet” and “complement”, whereas search for the keyword “coagulation” revealed no significant GO term. 7 GO terms were found by the keyword “lipoprotein”. GO terms with identical gene sets were not displayed. Detailed results and significantly regulated genes are listed in the Supplemental Table S8. Red z-scores indicate upregulated gene expression in SARS-CoV-2 infected cardiac tissue. G: 6 significant GO terms were identified for the keyword “MHC”. GO terms for MHC in general (1, 2) or more specified to MHC-I (3-6) were significantly regulated. MHC-II related GO terms were not significant. Detailed results and significantly regulated genes are listed in the Supplemental Table S9. Red z-scores indicate upregulated gene expression in SARS-CoV-2 infected cardiac tissue.

Second, the list of all 940 significant GO terms was searched for specific keywords. According to the current literature with respect to SARS-CoV-2, we chose keywords linked to four general topics: (i) virus, viral, (ii) lipoprotein, (iii) coagulation, platelet, complement, and (iv) MHC. The resulted GO terms with at least 10 annotated genes were depicted in Figure 6E-G.

As shown in Figure 6E, the search using the keywords “virus” and “viral” revealed 13 significant GO terms, which are linked to the general topic response to virus. As depicted in the outer circle, these GO terms include more genes significantly upregulated in SARS-CoV-2 infected cardiac tissue (red) than significantly downregulated genes (blue) compared to non-infected cardiac tissue. The ubiquitous upregulation of these GO terms is also demonstrated by red z-scores in the inner circle.

In Figure 6F, the keyword “lipoprotein” revealed seven significant GO terms, including four gene sets comprised of two identical sets. Therefore, five significant GO terms were displayed including the more specified GO terms for high-density (5) and very low density (9) lipoprotein particles. The keyword “platelet” revealed three significant GO terms (1-3), the keyword “complement” one significant GO term (4) and the keyword “coagulation” revealed no significant GO terms.

In Figure 6G, significant GO terms resulting from the keyword “MHC” were displayed. Remarkably, only GO terms with respect to MHC in general (1, 2) or more specified to MHC-I (3-6) were significantly regulated, whereas MHC-II related GO terms were not significant.

## 5 Discussion

Reporting on the largest available autopsy series so far, this analysis confirms that cardiac tissue can be infected by SARS-CoV-2 with virus replication being present in COVID-19 autopsy cases.^11^ Beyond this, the key findings of the study are: (i) virus replication in the myocardium is associated with accelerated death; (ii) expression of the virus entry factor transmembrane protease serine 2 (*TMPRSS2*) was increased in cases with cardiac SARS-CoV-2 infection while the *ACE2* receptor was not differently regulated and (iii) bulk mRNA sequencing revealed 824 differentially expressed genes in cases with cardiac infection followed by GO enrichment analyses allowing functional interpretations.

### 5.1 SARS-CoV-2 infection of the heart – susceptibility and consequences

This cohort study was performed in the metropolitan area of Hamburg, Germany. 85% of the deceased were aged 70 years or older, which is consistent with COVID-19 deaths in Germany.^27^ Virus infection of non-respiratory tract organs was documented in COVID-19 autopsy studies earlier.^5, 8, 28^ However, we recently showed that SARS-CoV-2 can be detected in the heart of about 50% of fatal COVID-19 cases.^11^ By analyzing more cases, we confirmed that the heart is targeted by SARS-CoV-2 in approximately 40-50% of the COVID-19 autopsy cases.

Next, we investigated the localization of virus particles within distinct cell types in the heart. In some studies, electron microcopy was utilized to confirm virus localization in the myocardial tissue.^29, 30^ However, the identification of virus particles by electron microscopy is unspecific and remains challenging, particularly in tissue sections.^31, 32^ Therefore, we combined two sequence-specific methods to confirm virus in the myocardium: (i) sequence-specific RT-PCR to determine virus presence and (ii) sequence-specific in situ hybridization for visualization and localization. Furthermore, fluorescent in situ hybridization revealed viral RNA in the interstitium or interstitial cells, whereas viral RNA in cardiomyocytes was rarely detectable but not absent. This created evidence that non-myocytes but also myocytes are targeted by the virus, coherent to other virus infections that do not target primarily myocytes and still cause myocardial dysfunction.^33, 34^

It is unclear which factors determine the susceptibility of the myocardium to be infected with SARS-CoV-2. Therefore, gene expression of *ACE2*, known as receptor of the Spike protein, and the host proteases *TMPRSS2, CTSB*, and *CTSL* was investigated.^35-37^ Interestingly, the infection of myocardial cells was associated with increased gene expression of the transmembrane protease serine 2 (*TMPRSS2*) in those cases with SARS-CoV-2 genome in the cardiac tissue and amplified in those with signs of virus replication. TMPRSS2 is facilitating virus entry into the host cell as well as being essential for virus spread and pathogenicity.^38^ This is the first report of *TMPRSS2* being differently regulated within cardiac tissue and therefore potentially mediating virus infection of cardiac tissue. Together, this might fuel therapeutic application of TRMPSS2 inhibitors for COVID-19, as are being tested in patients in clinical trials (NCT04321096 and NCT04338906). Obviously, further studies need to verify this finding, but intriguingly, TMPRSS2 knock-out reduced the inflammatory response after experimental SARS-CoV-2 infection and this was in part dependent on TLR3 signaling.^39^ This is coherent to our data showing less inflammatory dysregulation (gene expression of proinflammatory cytokines) in those without SARS-CoV-2 infection in the heart. Contrarily, *ACE2*, as the main receptor for virus uptake, as well as was the proteases cathepsin L and B (*CTSL*/*CTSB*), known to be involved in viral membrane fusion^35^ and priming the Spike protein of the virus, were not dysregulated in those groups, suggesting that susceptibility to the virus might not be correlated to changes in gene expression of those targets.

Additionally, relevant virus replication in the myocardium was detected in 15% of deceased. Neither virus infection nor virus replication in the heart was associated with increased numbers of inflammatory cells. This was coherent for different inflammatory cell markers including different T-cell subtypes as well as macrophages. Case reports with positive cardiac SARS-CoV-2 infection together with histological signs of myocarditis are rare,^40, 41^ whereas myocardial inflammation of patients recovered from COVID-19 but without confirmed SARS-CoV-2 infection in the heart was reported.^7^ Therefore, myocarditis might be developed as sequela due to active or prior COVID-19 disease which does not necessarily include cardiac infection with SARS-CoV-2. Whether myocardial virus activity in the absence of clinical evident myocarditis might result in long-term consequence is unknown.

In our study, 93% of cases suffered from two or more comorbidities which is coherent with other autopsy studies beyond Hamburg.^28, 42-44^ Interestingly, neither cardiac nor other comorbidities were associated with cardiac infection and even signs of virus replication in the myocardium were not more frequent in those with comorbidities. This is different in other organs, like the kidney, where cases with organ manifestation in general were more likely to have more comorbidities.^5, 8^ Age might also be a predictor to fuel organotropism of SARS-CoV-2, but all cases included here were of high age making this difficult to assess. In general, autopsy studies need to be interpreted with caution in view of these associations, since survivors of the disease are excluded, which makes this subgroup of COVID-19 patients unique. Despite that fact, virus replication in the heart is associated with more rapid death compared to those deceased without cardiac virus infection in our study cohort, suggesting that active cardiac infection could be a contributor to death. This is a clinically important and novel finding. Similarly, SARS-CoV-2 infection of kidney resulted in premature death as well.^5^

### 5.2 RNA sequencing and functional GO enrichment analyses

To address virus associated alterations in gene expression, we measured these changes utilizing bulk mRNA sequencing. Therefore, 10 cases with and 10 cases without cardiac SARS-CoV-2 infection were analyzed to detect differentially expressed genes. Thus, 824 significantly regulated genes (p < 0.05) were identified suggesting these might be specific for cardiac SARS-CoV-2 infection. Out of the identified 824 genes, 73 proteins were measured in plasma of a COVID-19 cohort by Olink proteomics.^26^ The SARS-CoV-2 associated regulation on gene expression level within the myocardium was confirmed for 17 plasma proteins (16 upregulated, 1 downregulated) in severely diseased patients with and without COVID-19.

Applying a FDR < 0.05, the number of 824 differentially expressed genes were subsequently narrowed down to the top 19 genes. Interestingly, those contained two upregulated apolipoproteins (*APOE* and *APOD*) and two upregulated interferon related proteins (*IFI44L* and *IFIT3*) as well as upregulated *TRIM25* known as interferon expression inducing factor. Wang et al. previously showed that ACE2 as virus receptor was translocated into lipid rafts as viral entry site. This translocation can be induced by APOE-mediated cholesterol influx. Using a SARS-CoV-2 pseudovirus entry assay, they documented a 3- fold higher virus infection in cells treated with APOE.^45^ Upregulated APOD expression was shown during acute encephalitis induced by the human coronavirus OC43 (HCoVC-OC43) in mice, while overexpressing *ApoD* in transgenic mice resulted in improved survival.^46^

Interferons are central to antiviral immunity, but impaired interferon activity was described in severe COVID-19 patients.^47^ In line with this finding, our second top gene *IFI44L* was described as negative feedback regulator of host antiviral response. *IFI44L* silenced cells revealed reduced virus titer indicting that IFI44L supports virus replication.^48^

To go beyond single genes, GO enrichment analyses were performed resulting in 940 significant GO terms all together. To identify relevant GO terms, we utilized 3 different approaches to narrow down this large number of GO terms. First, GO terms including at least three of the top 19 differentially expressed genes were determined and plotted as GO chord graph. Second, most significant child terms were determined according to ancestor chars to avoid lists of child and parent terms. Third, besides those untargeted selection, keywords were used to filter significant GO terms according to virus associated host response, such as MHC molecules, or clinically known features, like dyslipidemia or thrombosis.^49, 50^

The SARS-CoV-2 infection elicits typical transcriptional response in the cardiac tissue. This includes for instance the GO terms “defense response to virus”, “innate immune response” and “type I interferon signaling pathway”. Interestingly, the GO term “defense response to virus” was identified by both untargeted approaches strengthening the specificity of this term. Similarly, the GO term “extracellular exosomes” were also identified by both untargeted selection criteria. This unexpected GO term represents the most significant GO term comparing all descendent child terms. This is in line with the finding that the virus life cycle shares common routes with the biogenesis and release of extracellular vesicles. Additionally, extracellular vesicles are involved in immune response against viral pathogens by incorporating and spread both viral and host factors. There is evidence that viruses hijack the secretory pathway for extracellular vesicles to exit infected cells.^51^ Interestingly, during virus infection, these extracellular vesicles may become a delivery vector of viral material. This is generally thought to be a pro-viral effect and might enable spreading the virus also within the heart.

Furthermore, in SARS-CoV-2 positive cardiac tissue decreased gene expression (indicated by blue z-scores) were found in GO terms annotated for cardiomyocyte structural proteins. This is in line with the transcriptomic finding previously reported for SARS-CoV-2 infected human iPSC-derived cardiomyocytes indicting cytopathic effects on cardiomyocytes.^52^

Utilizing the third approach to select relevant GO terms by specific keywords, all significant GO terms related to virus are upregulated in SARS-CoV-2 infected cardiac tissue indicated by red z-scores. This documents the response of myocardial cells to SARS-CoV-2 infection. Next, the keyword “MHC” revealed significant GO terms annotating MHC complexes in general and more specified MHC-I class, which are important for presenting viral proteins of infected cells. The recognition of viral peptides complexed with MHC-I molecules is one key event to eliminate virus-infected cells.^53^

Vascular thromboembolisms is frequent in COVID-19 patients treated in an intensive care setting.^49, 54^ Therefore, we extended the keyword-based selection to known clinical features such as thrombosis and dyslipidemia. Therefore, the keywords “platelet”, “complement” and “coagulation” as well as “lipoproteins” were applied to identify significant GO terms. While the keyword coagulation did not reveal significant GO terms, platelet and complement resulted in upregulated GO terms as indicated by red z-scores.

Besides the fact that two apolipoproteins were already identified within the top 19 differentially expressed genes, the keyword lipoprotein revealed significant GO terms upregulated in SARS-CoV-2 infected cardiac tissue. Several studies proposed an association between dyslipidemia and the severity of COVID-19,^55^ which is further strengthened by our transcriptomic findings.

## 6 Limitation

This study has some limitations, including the design as an autopsy study. Elderly age and frequently observed comorbidities of deceased might have influenced the result of cardiac infections and replication. Presumably, in non-autopsy studies, the incidence of virus infection will be lower. Unfortunately, endomyocardial biopsies of patients with COVID-19 are rare and especially no clinical routine so far. As an autopsy study, we have only limited clinical information and no information is present about myocardial biomarkers, which might be upregulated due to the SARS-CoV-2 infection or to systemic inflammation due to COVID-19. Unfortunately, post mortem biochemistry of cardiac biomarkers in cadaveric serum is not useful even within the first 48 h after death.^56^ The presented transcriptomic data provide a snapshot at the time of autopsy, whether differentially expressed genes are the cause or a consequence of cardiac infection needs to be investigated in the future. Since all cases were tested positive for SARS-CoV-2 infection, we cannot identify factors mediating an impaired cardiac function due to systemic inflammation caused by COVID-19.

## 7 Conclusion

Taken together, SARS-CoV-2 seems to alter transcriptomic pathways after infecting the heart but not causes myocarditis in deceased died with COVID-19. Nevertheless, this was associated with accelerated death of cases with vs. without cardiac replication. Elucidating the immune mechanisms of cardiac involvement as well as other factors contributing to disease severity will provide further insights into early and late subsequent virus-associated myocardial damage. Moreover, increasing pathophysiological understanding might allow for precise and targeted interventions.

## Data Availability

Please contact corresponding authors for data request.

## Acknowledgments

We thank the UKE Microscopy Imaging Facility (Umif), University Hospital Center Hamburg-Eppendorf for providing microscopes and support, GenXPro for performing MACE analyses and bioinformatics. Data for plasma protein levels were provided by the MGH Emergency Department COVID-19 Cohort (Filbin, Goldberg, Hacohen) with Olink Proteomics. Figure 1 was created with BioRender.com.

## Funding

This work is supported by the German Centre for Cardiovascular Research (DZHK) (FKZ 81Z0710108 to D.W.), by the grant to D.L. and D.W. from the Deutsche Herzstiftung and by CRC1192 to TBH.

## Disclosures

Dr Blankenberg has received research funding from Abbott Diagnostics, Bayer, SIEMENS, Singulex and Thermo Fisher. He received honoraria for lectures from Abbott, Abbott Diagnostics, Astra Zeneca, Bayer, AMGEN, Medtronic, Pfizer, Roche, SIEMENS Diagnostics, SIEMENS, Thermo Fisher and as member of Advisory Boards and for consulting for Bayer, Novartis and Thermo Fisher unrelated to the submitted work. Dr Kirchhof reports research support for basic, translational, and clinical research projects from European Union, British Heart Foundation, Leducq Foundation, Medical Research Council (United Kingdom), and German Center for Cardiovascular Research; support from several drug and device companies active in atrial fibrillation; and has received honoraria from several such companies in the past, but not in the past 3 years. He is listed as inventor on 2 patents held by the University of Birmingham (Atrial Fibrillation Therapy, WO 2015140571; Markers for Atrial Fibrillation, WO 2016012783), unrelated to the submitted work. Dr Kluge received research support from Ambu, Daiichi Sankyo, ETView Ltd, Fisher & Paykel, Pfizer and Xenios. He also received lecture fees from Astra, Basilea, C.R. Bard, Baxter, Biotest, Cytosorbents, Fresenius, Gilead, MSD, Pfizer, Philips and Zoll. He received consultant fees from Baxter, Fresenius, Gilead, MSD and Pfizer. Dr Westermann reports receiving a speaker’s fee from AstraZeneca, Bayer, and Novartis, unrelated to the submitted work.

## Notes

### Author Declarations

This study was approved by the local ethics committee of the Hamburg Chamber of Physicians (PV7311).

