## Supplemental Data for "Cardiac SARS-CoV-2 infection is associated with distinct transcriptomic changes within the heart"

[1] Department of Cardiology, University Heart and Vascular Center Hamburg, Germany.
[2] DZHK (German Center for Cardiovascular Research), partner site Hamburg/Kiel/Lübeck.
[3] Institute of Legal Medicine, University Medical Center Hamburg-Eppendorf, Germany.
[4] Institute for Cardiac Diagnostics and Therapy, Berlin, Germany.
[5] Department of Electron Microscopy, Center for Molecular Neurobiology, University Medical Center Hamburg-Eppendorf, Germany.
[6] Department of Cardiology, Charité-Universitaetsmedizin, Berlin, Germany.
[7] DZHK (German Centre for Cardiovascular Research), partner site Berlin, Germany.
[8] III. Department of Medicine, University Medical Center Hamburg-Eppendorf, Germany.
[9] GenXPro GmbH, Frankfurter Innovationszentrum, Biotechnologie (FIZ), Frankfurt am Main, Germany.
[10] Department of Intensive Care Medicine, University Medical Center Hamburg-Eppendorf, Germany.
[11] Institute of Cardiovascular Sciences, University of Birmingham, UK.
[12] Division of Cardiology (cNEP), Evangelisches Krankenhaus Düsseldorf, Germany.
[13] Institute of Neural and Sensory Physiology, Medical Faculty, Heinrich Heine University Düsseldorf, Germany.
* these authors contributed equally to this work.

**Corresponding authors**
✉ Dirk Westermann, MD


✉ Diana Lindner, PhD


University Heart and Vascular Center Hamburg
Department of Cardiology
Martinistr. 52, 20246 Hamburg, Germany

### 1 Expanded Material and Methods

#### 1.1 RNA isolation and reverse transcription

To isolate total RNA, snap frozen cardiac tissue was added to QIAzol (Qiagen, Germany) and immediately disrupted in the tissue lyser II (Qiagen, Germany). Using the miRNeasy mini kit (Qiagen, Germany), RNA was further purified following the manufacturer's protocol. As suggested, DNase I (Qiagen, Germany) was applied to the column to avoid genomic DNA contamination. RNA concentration was measured on a Nanodrop 2000c spectrophotometer. Until further processing, RNA was stored at -80 °C. The synthesized complementary DNA (cDNA) was diluted to a final concentration of 10 ng/µl prior to TaqMan analyses.

#### 1.2 Gene expression analysis – TaqMan PCR

Quantitative TaqMan PCR was performed on a QuantStudio7 system (Thermo Fisher Scientific, USA) with 2.5 µl gene expression master mix (Thermo Fisher Scientific, USA) and 0.25 µl specific gene expression assay. Gene expression assays, including forward and reverse primers as well as the FAM-labelled probe, are listed in the Supplemental Table S1. cDNA samples were analyzed in duplicates. Due to the low gene expression of *ACE2* and *TMPRSS2*, some samples revealed no Ct value for these target genes. In these cases, the Ct value was set to 40 for subsequent calculations. The Ct value of the housekeeping gene *CDKN1b* was used to calculate ΔCt values of the target genes. The median ΔCt value of the group without cardiac infection was used to quantify the gene expression using the formula $2^{-\Delta\Delta Ct}$ and plotted as relative gene expression x-fold to "no cardiac infection".

**Supplemental Table S1: Gene expression assays purchased from Thermo Fisher Scientific.**

| Assay | Gene name | Ordering number |
|---|---|---|
| ***ACE2*** | Angiotensin-converting enzyme 2 | Hs00222343_m1 |
| ***TMPRSS2*** | Transmembrane protease serine 2 | Hs01122322_m1 |
| ***CTSB*** | Cathepsin B | Hs00947439_m1 |
| ***CTSL*** | Cathepsin L | Hs00964650_m1 |
| ***CDKN1b*** | Cyclin-dependent kinase inhibitor 1B | Hs00153277_m1 |

#### 1.3 Histological analyses

Left ventricular tissue was fixed in 10% neutral-buffered formalin for at least 48 h at room temperature (RT), subsequently dehydrated and embedded in paraffin. Four micrometer-thick sections were cut and deparaffinized using xylene substitute (Roti-Histol, Germany) and rehydrated in a descending ethanol series. After dehydration, slides were mounted in Eukitt (Sigma-Aldrich, USA).

To characterize and quantify inflammatory infiltrates, FFPE tissue sections underwent a heat-induced antigen retrieval using citrate butter (pH 6). For detection of immune cell-specific markers, the primary antibodies used are listed in the Supplemental Table S2. As secondary antibody, we used enhancing EnVision™ peroxidase-conjugated anti-mouse antibody (DakoCytomation, Germany). Immunohistological staining was visualized using 3-amino-9-ethylcarbazole (Merck, Germany) as chromogenic substrate. Finally, slides were counterstained in haematoxylin and mounted with Kaiser's gelatine (Merck, Germany). Immunoreactivity was quantified by digital image analysis (DIA) as described previously.[1]

**Supplemental Table S2: Primary antibodies used for immunohistological or immunofluorescence analyses.**

| Antibody | Species | Dilution | Company | Ordering information |
|---|---|---|---|---|
| Primary antibodies | | | | |
| anti- human CD3 | mouse | 1:25 | Agilent | M7254 |
| anti- CD8 | mouse | 1:50 | Medac | 108M-94 |
| anti- human CD68 | mouse | 1:100 | Agilent | M0876 |
| anti- human CD45R0 | mouse | 1:300 | Agilent | M0742 |
| anti- $\alpha$- actinin | rabbit | 1:30 | Cell Signaling | 3134 |
| Secondary antibodies | | | | |
| Anti- rabbit - alexafluor- 488 | donkey | 1:500 | Thermo Fisher | A-21206 |

In case of fluorescence staining, heat-induced antigen retrieval was performed using citrate buffer (pH 6). Subsequently, sections were permeabilized in 0.2% Triton X-100/tris-buffered saline (TBS) for 10 min and autofluorescence was quenched with the help of 0.25% Sudan black/70% ethanol for 30 min. Before application of primary antibodies (see Supplemental Table S2) over night at 4 °C, sections were blocked with 3% bovine serum albumin (BSA)/TBS for 2 h at RT. The secondary antibody listed in the Supplemental Table S2 was incubated for 2 h RT. After washing in TBS, slides were mounted in DAPI Fluoromount-G (SouthernBiotech, USA).

#### 1.4 RNA in situ hybridization

For detection of SARS-CoV-2 plus and minus strand, RNAscope in situ hybridization was performed using chromogenic staining or fluorescent staining on FFPE tissue sections. For chromogenic staining, RNAscope® 2.5 HD Reagent Kit-RED (#322350; Advanced Cell Diagnostics (ACD), USA) was used, whereas for localization studies, RNAscope Multiplex Fluorescent v2 reagent kit (#323100, ACD, USA) was used. After deparaffinization, target retrieval was performed for 15 min, followed by quenching of internal peroxidase activity and permeabilization by protease plus treatment for 30 min at 40°C. The hybridization of target specific probe (see Supplemental Table S3), subsequent amplification steps and signal detection were performed as described in manufacturer's instructions. For chromogenic staining, nuclei were counterstained using hematoxylin. In case of fluorescent staining, lipofuscin autofluorescence was quenched via Sudan black staining (0.25% in 70% ethanol) for 8 min, followed by a blocking step (1% BSA/TBS for 1 h at RT) and subsequent counterstaining with Alexa-coupled wheat germ agglutinin (WGA, 1:500) and bisbenzimide H33342 trihydrochloride (Hoechst, 1:1000) for 1 h in blocking solution. Matching negative controls for chromogenic and fluorescent staining are shown in the Supplemental Figures S1 and S2, respectively.

**Supplemental Table S3: Probes used for in situ hybridization (RNAscope).**

| Probe | Company | Ordering information |
|---|---|---|
| RNAscope® Probe- V-nCoV2019-S | ACD | 848561 |
| RNAscope® Probe- V-nCoV2019-S-sense | ACD | 845701 |
| RNAscope® Positive Control Probe- Hs-UBC | ACD | 310041 |
| RNAscope® Negative Control Probe- DapB | ACD | 310043 |

#### 1.5 Microscopy

Images of non-fluorescently stained tissue including chromogenic RNAscope staining were taken using a Keyence BX-9000 microscope equipped with ×20 plan APO (NA = 0.75) and ×60 plan APO oil (NA = 1.40) objectives. After immunofluorescence staining and fluorescent RNAscope, confocal images were captured using a Leica TCS SP5 confocal microscope (Leica Microsystems) equipped with ×40 HCX PL APO CS oil (NA = 1.3) and ×63 HCX PL APO Lbd. Bl. oil (NA = 1.4-0.6) objectives. Three-dimensional images were collected and a maximum projection image was created using the Leica LAS AF software.

### 2 Expanded Results

#### 2.1 Expanded baseline information

The extended baseline table (Supplemental Table S4) provides further information concerning case characteristics and data availability. The added category, location of death, is grouped in four classifications: domesticity, nursing home, normal care, and intensive care. Furthermore, all characteristics for the groups analyzed by 3’mRNA sequencing are listed. To improve RNA quality for 3’mRNA sequencing, patients died in hospital and with post mortem intervals of maximal five days were chosen. However, the cases represent the entire study cohort regarding the categories age, sex, time between diagnosis and death, comorbidities and BMI.

**Supplemental Table S4: Extended baseline characteristics.**

| | | Cases grouped by cardiac virus load or replication | | | | | Cases analyzed by 3'mRNA sequencing | | |
|---|---|---|---|---|---|---|---|---|---|
| | **All cases** (N=95) | No cardiac infection (N=46) | Cardiac infection (> 1000 copies) (N=41) | p-value* | Virus replication (scored >1) (N=14) | p-value* | No cardiac infection (N=10) | Cardiac infection (>1000 copies) (N=10) | p-value† |
| **Female No. (%)** | 51 (53.7) | 21 (45.7) | 24 (58.5) | 0.32 | 7 (50.0) | 1.00 | 5 (50.0) | 7 (70.0) | 0.65 |
| Available data No. (%) | 95 (100) | 46 (100) | 41 (100) | | 14 (100) | | 10 (100) | 10 (100) | |
| **Age [years]** | 84.0 (76.0, 87.0) | 81.5 (74.2, 86.0) | 84.0 (78.0, 88.0) | 0.27 | 86.0 (82.5, 87.8) | 0.13 | 82.5 (78.2, 84.0) | 84.5 (79.8, 86.5) | 0.34 |
| Available data No. (%) | 95 (100) | 46 (100) | 41 (100) | | 14 (100) | | 10 (100) | 10 (100) | |
| **Post mortem interval [days]** | 4.0 (3.0, 7.0) | 4.0 (2.0, 6.0) | 4.0 (3.0, 8.0) | 0.38 | 5.0 (3.0, 9.8) | 0.17 | 4.0 (1.5, 4.0) | 3.5 (2.2, 4.8) | 0.79 |
| Available data No. (%) | 93 (97.9) | 45 (97.8) | 40 (97.6) | | 14 (100) | | 10 (100) | 10 (100) | |
| **Location of death:** | | | | | | | | | |
| **Domesticity No. (%)** | 10 (11.1) | 7 (15.9) | 3 (7.9) | 0.44 | 0 (0) | 0.29 | 0 (0) | 0 (0) | |
| **Nursing home No. (%)** | 24 (26.7) | 7 (15.9) | 12 (31.6) | 0.16 | 5 (38.5) | 0.17 | 0 (0) | 0 (0) | |
| **Normal Care No. (%)** | 32 (35.6) | 15 (34.1) | 16 (42.1) | 0.60 | 8 (61.5) | 0.15 | 5 (50.0) | 8 (80.0) | 0.35 |
| **Intensive care No. (%)** | 23 (25.6) | 14 (31.8) | 7 (18.4) | 0.26 | 0 (0) | 0.048 | 5 (50.0) | 2 (20.0) | 0.35 |
| Available data No. (%) | 90 (94.7) | 44 (95.7) | 38 (92.7) | | 13 (92.9) | | 100 (100) | 100 (100) | |
| **Time, diagnosis until death [days]** | 9.0 (3.0, 18.0) | 12.5 (7.2, 20.2) | 8.0 (2.0, 10.0) | 0.082 | 2.0 (2.0, 9.0) | 0.017 | 12.0 (5.5, 19.5) | 8.0 (2.0, 10.0) | 0.43 |
| Available data No. (%) | 55 (57.9) | 26 (56.5) | 25 (61.0) | | 9 (64.3) | | 7 (70.0) | 9 (90.0) | |
| **Comorbidities** | | | | | | | | | |
| HTN No. (%) | 44 (46.3) | 24 (52.2) | 17 (41.5) | 0.43 | 6 (42.9) | 0.76 | 4 (40.0) | 5 (50.0) | 1.00 |
| CAD No. (%) | 58 (61.1) | 31 (67.4) | 22 (53.7) | 0.28 | 7 (50.0) | 0.39 | 5 (50.0) | 4 (40.0) | 1.00 |
| HF No. (%) | 49 (51.6) | 23 (50.0) | 22 (53.7) | 0.90 | 8 (57.1) | 0.87 | 4 (40.0) | 5 (50.0) | 1.00 |
| **Sum of cardiac comorbidities ≥2** | 48 (50.5) | 26 (56.5) | 18 (43.9) | 0.34 | 8 (57.1) | 1.00 | 2 (20.0) | 4 (40.0) | 0.63 |
| DM No. (%) | 18 (18.9) | 11 (23.9) | 5 (12.2) | 0.26 | 4 (28.6) | 1.00 | 3 (30.0) | 1 (10.0) | 0.58 |
| Stroke No. (%) | 16 (16.8) | 7 (15.2) | 7 (17.1) | 1.00 | 5 (35.7) | 0.19 | 2 (20.0) | 1 (10.0) | 1.00 |
| COPD No. (%) | 49 (51.6) | 23 (50.0) | 24 (58.5) | 0.56 | 10 (71.4) | 0.27 | 2 (20.0) | 5 (50.0) | 0.35 |
| Cancer No. (%) | 29 (30.5) | 13 (28.3) | 14 (34.1) | 0.72 | 3 (21.4) | 0.87 | 3 (30.0) | 7 (70.0) | 0.18 |
| Renal failure No. (%) | 34 (35.8) | 16 (34.8) | 17 (41.5) | 0.67 | 8 (57.1) | 0.24 | 2 (20.0) | 4 (40.0) | 0.63 |
| **Sum of non-cardiac comorbidities ≥2** | 48 (50.5) | 22 (47.8) | 23 (56.1) | 0.58 | 11 (78.6) | 0.086 | 3 (30.0) | 7 (70.0) | 0.18 |
| **Sum of comorbidities ≥2** | 88 (92.6) | 42 (91.3) | 38 (92.7) | 1.00 | 14 (100) | 0.60 | 8 (80.0) | 10 (100) | 0.46 |
| Available data No. (%) | 95 (100) | 46 (100) | 41 (100) | | 14 (100) | | 10 (100) | 10 (100) | |
| **BMI [kg/m²]** | 24.4 (20.7, 28.7) | 24.4 (20.9, 28.4) | 23.2 (19.1, 28.7) | 0.58 | 25.0 (21.6, 26.8) | 0.85 | 27.4 (25.2, 28.6) | 22.4 (20.9, 31.7) | 0.57 |
| Available data No. (%) | 84 (88.4) | 38 (82.6) | 39 (95.1) | | 13 (92.9) | | 9 (90.0) | 10 (100) | |

| | | | | | | | | | |
|---|---|---|---|---|---|---|---|---|---|
| **Heart weight [g]** | 450.0 (361.2, 520.0) | 455.0 (383.8, 510.0) | 420.0 (333.8, 530.0) | 0.40 | 410.0 (330.0, 505.0) | 0.27 | 452.5 (407.5, 517.5) | 460.0 (347.5, 533.8) | 0.76 |
| Available data No. (%) | 94 (98.9) | 46 (100) | 40 (97.6) | | 13 (92.9) | | 10 (100) | 10 (100) | |

For continuous variables median ($25^{th}$ percentile, $75^{th}$ percentile) are given. For binary variables absolute numbers and relative frequencies (%) are given. The presented p-values were determined by the comparison to all cases with no cardiac infection (N=46)[*] or to the 10 cases with no cardiac infection used for 3’mRNA sequencing (N=10)[†]. Mann-Whitney test was used for continuous variables and chi-squared test for binary variables. Abbreviations: BMI, body mass index; CAD, coronary artery disease; COPD, chronic obstructive pulmonary disease; DM, diabetes mellitus; HF, heart failure; HTN, hypertension; No, numbers.

#### 2.2 Controls for in situ hybridization

All tissue sections used for in situ hybridization underwent positive and negative controls. As positive control, a probe detecting the mRNA of the ubiquitously expressed Ubiquitin C was used, whereas a probe detecting RNA of the bacterial protein dihydro-dipicolinate reductase (DapB) was used as negative control. The positive control confirmed that the tissue quality enables the detection of RNA, the negative control demonstrate the specificity of the detected signals. Supplemental Figure S1 displays the positive and negative control for case - 08 displayed in Figure 2C in the main manuscript. Supplemental Figure S2 displays the positive and negative controls for the cases used for fluorescent staining depicted in Figure 3 in the main manuscript.

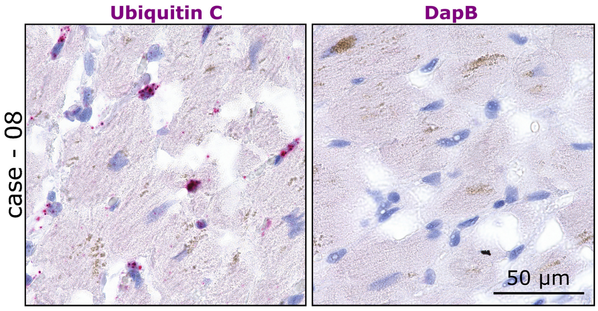


**Supplemental Figure S1: Controls for chromogenic RNAscope in situ hybridization.**
As positive control of successful chromogenic RNAscope in situ hybridization, probe against Ubiquitin C was used, revealing strong red signals in cardiac tissue. A probe detecting the bacterial gene dihydro-dipicolinate reductase (DapB) was used as negative control and revealed no specific staining.

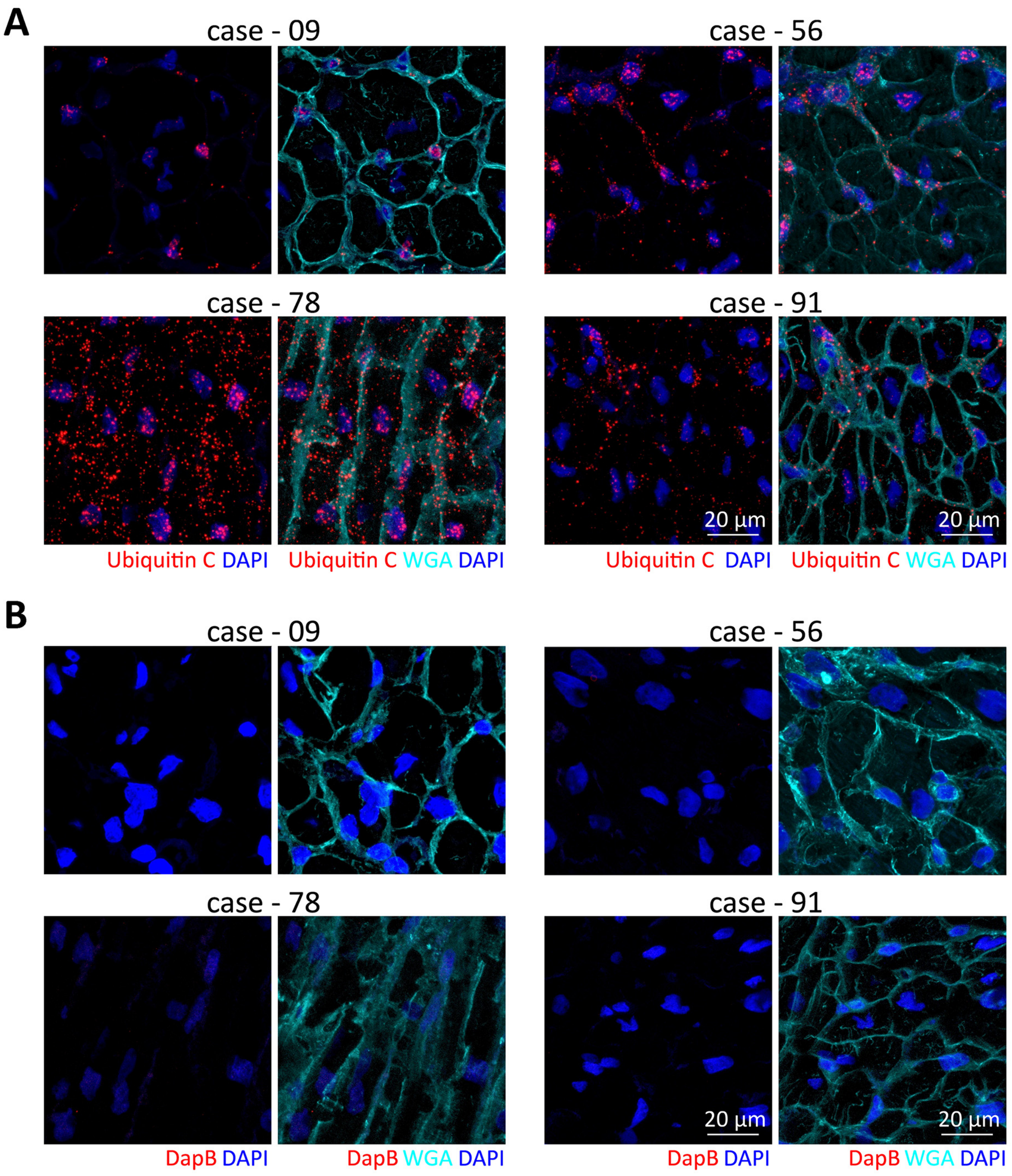


**Supplemental Figure S2: Controls for fluorescent RNAscope in situ hybridization.**
A: As positive control of fluorescent RNAscope in situ hybridization Ubiquitin C (red) was used, wheat germ agglutinin (WGA, cyan) was used to counterstain sections.
B: As negative control the bacterial gene dihydro-dipicolinate reductase (DapB, red) was applied on tissue sections. Cell membranes were counterstained with WGA (cyan).

#### 2.3 Olink Data

Olink proteomics performed measurements of 1429 proteins in plasma samples in a cohort of 384 patients including 306 SARS-CoV-2 infected patients and 78 symptomatic controls.[2] Data were provided by the MGH Emergency Department COVID-19 Cohort (Filbin, Goldberg, Hacohen) with Olink Proteomics. In the present study 824 differently expressed genes in the SARS-CoV-2 infected myocardium were detected. Those were aligned with the 1429 proteins measured by Olink proteomics, resulting in 73 overlapping genes and proteins (Supplemental Figure S3A). The aim of the subsequent analysis was to compare the regulation of these 73 targets on plasma protein and myocardial gene expression level.

Out of all patients included in Olink proteomics analysis, two groups were formed for further analysis (Supplemental Figure S3B). Thus, in the first step all patients with pre-existing heart disease were excluded. Next, all patients classified in WHO category 2 (intubated, ventilated) on day 0 were selected and divided in patients with and without COVID-19. This results in two groups with same disease severity to uncover COVID-19-specific regulated markers.

The plasma protein expression on day 0 of the Olink proteomic study was compared between those groups. Out of the 73 plasma proteins, 20 were upregulated and 3 were downregulated. Overlaps with the significantly up- or downregulated genes were displayed as Venn diagram (Supplemental Figure S3C). Out of the 48 myocardial upregulated genes, 16 were confirmed to be elevated on plasma protein level. However, for downregulated genes this accordance cannot be observed: From 24 downregulated genes in SARS-CoV-2 infected myocardium only one protein is downregulated on plasma protein level. The exact p-values and expression data of those 17 proteins are listed in Supplemental Figure S3D.

**A**

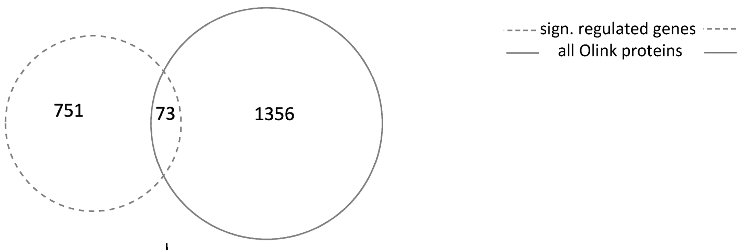


ADA2, AGER, APEX1, BGN, BPIFB1, BST2, BTN3A2, CA4, CCDC80, CCL20, CD177, CD34, CD48, CD63, CDHR1, CDKN1A, COL1A1, COX5B, CRNN, CSF3, CTSH, CXCL1, CXCL16, DAB2, DLK1, DNAJB1, EPCAM, EPHB4, FRZB, HLA-E, HPCAL1, HSPA1A, HSPB6, ICAM1, IGFBP4, IL1R2, IL33, IL6R, INPPL1, ITGB1BP2, KRT19, LAP3, LAT2, LCN2, LTBR, MB, MPHOSPH8, MSMB, NPPB, NRP2, PDCD5, PILRB, PRDX6, PRKRA, PRSS8, PSMA1, SLC16A1, SLC39A14, SMPD1, SPINK5, TIMP1, TNC, TNFRSF10A, TNFRSF14, TNFRSF1B, TNFSF10, TNFSF14, TYMP, USO1, VASH1, VEGFD, WARS, WFDC2

**B**

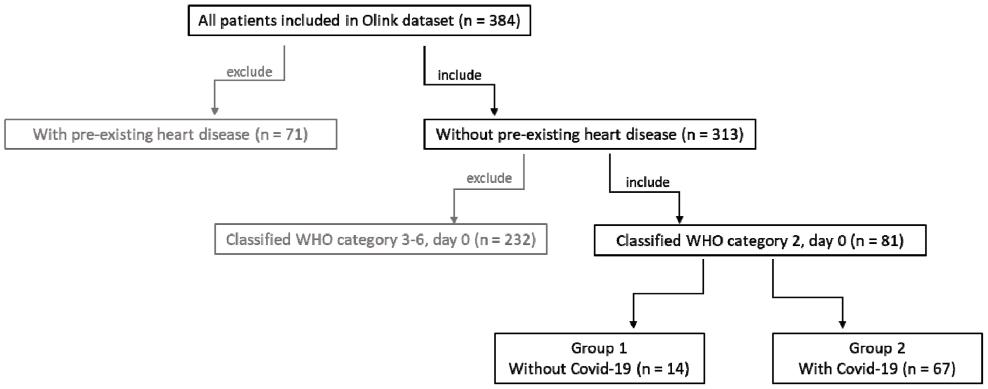


**C**

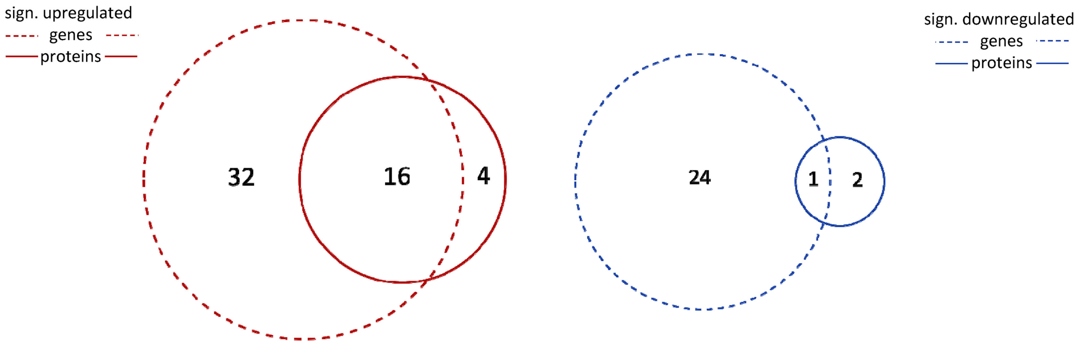


AGER, BST, BTN3A2, CCL20, CD177, CD48, CTSH, HPCAL1, HSPA1A, KRT19, LAP3, MPHOSPH8, TNFRSF10A, TNFSF14, TYMP, WARS

HSPB6

**D**

| Protein | Without Covid NPX (Median, IQR) | With Covid NPX (Median, IQR) | p-value |
|---|---|---|---|
| AGER | 5.1 (4.4, 5.3) | 6.4 (5.9, 8.4) | 0.0002 |
| BST2 | 3.2 (2.7, 3.9) | 5.0 (4.2, 5.9) | <0.0001 |
| BTN3A2 | 3.0 (2.6, 3.3) | 3.2 (2.9, 3.6) | 0.0415 |
| CCL20 | 5.2 (4.6, 5.4) | 5.9 (5.1, 6.7) | 0.0415 |
| CD177 | 3.2 (2.2, 3.9) | 3.7 (3.2, 4.3) | 0.0499 |
| CD48 | 4.3 (4.0, 4.5) | 4.7 (4.2, 5.1) | 0.0187 |
| CTSH | 3.0 (2.6, 3.3) | 3.6 (3.0, 4.1) | 0.0020 |
| HPCAL1 | 4.3 (3.2, 5.0) | 5.0 (4.2, 5.9) | 0.0207 |
| HSP1A1 | 2.0 (1.3, 2.8) | 2.4 (2.0, 3.8) | 0.0469 |

| Protein | Without Covid NPX (Median, IQR) | With Covid NPX (Median, IQR) | p-value |
|---|---|---|---|
| HSPB6 | 4.7 (3.8, 5.9) | 3.3 (2.7, 4.5) | 0.0042 |
| KRT19 | 2.3 (1.4, 3.1) | 4.7 (3.7, 5.5) | <0.0001 |
| LAP3 | 2.6 (2.4 ,3.1) | 5.0 (4.3, 5.6) | <0.0001 |
| MPHOSPH8 | 0.4 (0.1, 0.8) | 0.9 (0.6, 1.3) | 0.0173 |
| TNFRSF10A | 3.4 (2.6, 3.6) | 3.7 (3.3, 4.2) | 0.0425 |
| TNFSF14 | 2.8 (2.2, 3.1) | 3.7 (3.4, 4.1) | <0.0001 |
| TYMP | 3.8 (3.2, 5.1) | 6.0 (5.4, 6.6) | <0.0001 |
| WARS | 4.2 (3.9, 5.4) | 6.0 (5.5, 6.3) | <0.0001 |

**Supplemental Figure S3: Overlaps of significantly regulated genes with proteins measured in the Olink proteomics dataset.**

A: Due to SARS-CoV-2 infection of the myocardium, 824 differentially expressed genes were identified in this study and aligned with 1429 proteins measured in the Olink proteomics COVID-19 dataset. 73 overlapping genes and proteins were detected (abbreviations under parenthesis).

B: To identify SARS-CoV-2 specific plasma markers, two groups were formed out of 384 patients. Patients classified in WHO category 2 (intubated and ventilated) on day 0 were selected. These patients were divided in patients without COVID-19 (group 1) and with COVID-19 (group 2). Patients with pre-existing heart diseases were excluded.

C: In red: 48 genes and 20 proteins were identified as upregulated resulting in 16 overlaps upregulated on both gene expression and protein level. In blue: 25 genes and 3 proteins are downregulated resulting in only one overlap, downregulated on both gene expression and protein level.

D: The median and interquartile range (IQR) of the normalized protein expression (NPX) of the two groups defined in B are shown. With the gene expression overlapping proteins are listed. P-values were calculated using the Mann-Whitney test.

#### 2.4 Expanded data from Gene Ontology enrichment analyses

In the presented manuscript, 5 GO circles are displayed. Detailed information and a list of all significantly regulated genes are provided in the following Supplemental Tables S5 - S9.

For the GO circle displayed in Figure 6A, we aimed to present the overall most significant GO terms. But often the significance of parental and child terms is based on the same gene set. For example, the three most significant GO terms in the category biological process include nearly the similar set of genes all involved in interferon signaling (Supplemental Figure S4). Thus, to avoid a list of child terms including the same set of genes, the most specified GO terms within ancestor charts were selected. For this reason, ancestor charts of the 15 most significant GO terms per category were created. Data visualization of ancestor charts was provided by a GenXPro Web application at www.tools.genxpro.net. In the next step, the most descendent child terms including at least 40 annotated genes were selected. The ancestor charts for the three categories biological process, cellular component, and molecular function are depicted in the Supplemental Figures S4, S5, and S6, respectively. The identified GO terms are numbered according to their appearance in Figure 6A.

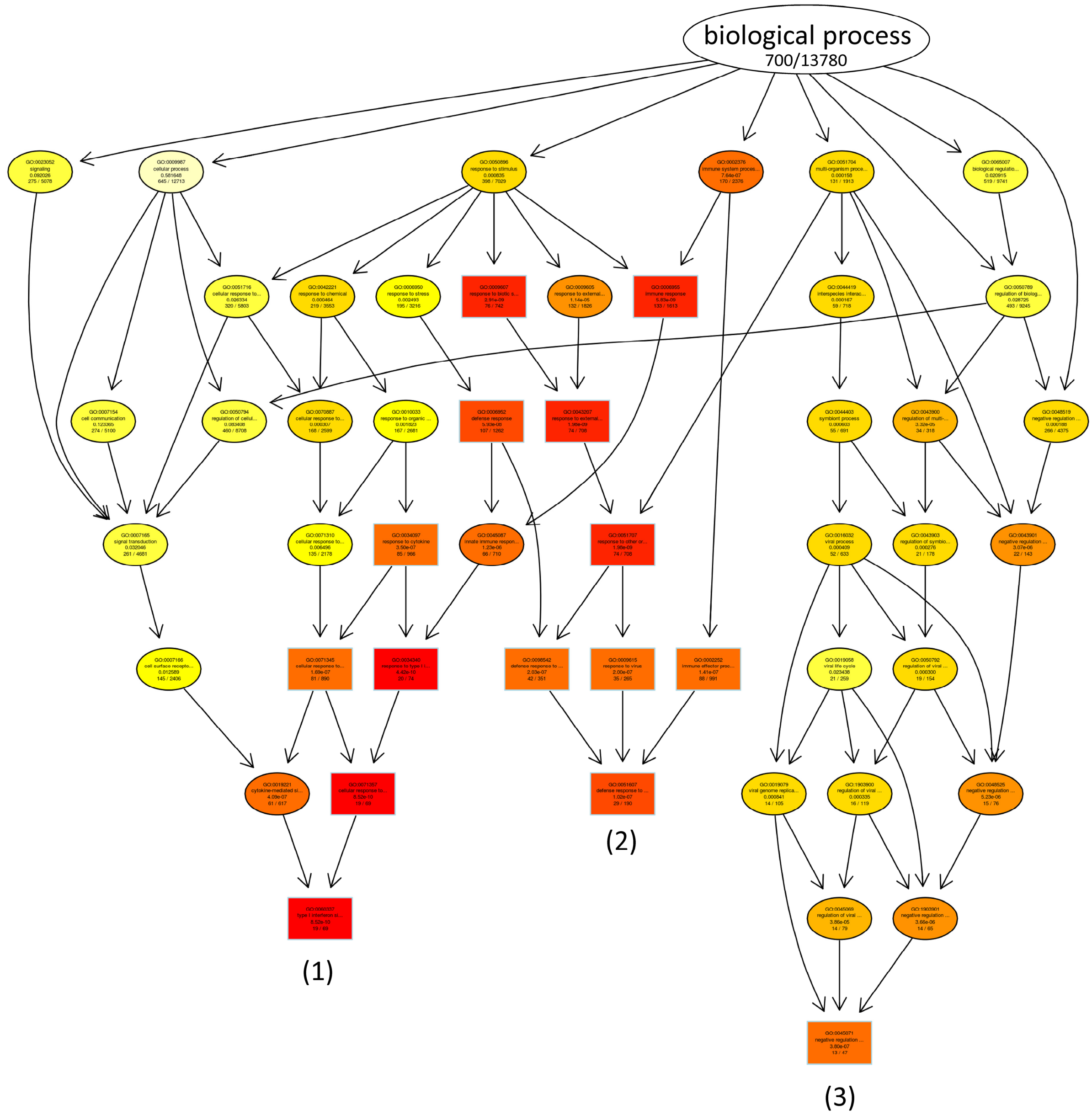


**Supplemental Figure S4: Ancestor Chart created for the 15 most significant GO terms in the category biological process.**

The 15 most significant GO terms were displayed as squares, the connected GO terms as circles. The color is defined by the significance illustrating the most significant terms in red.

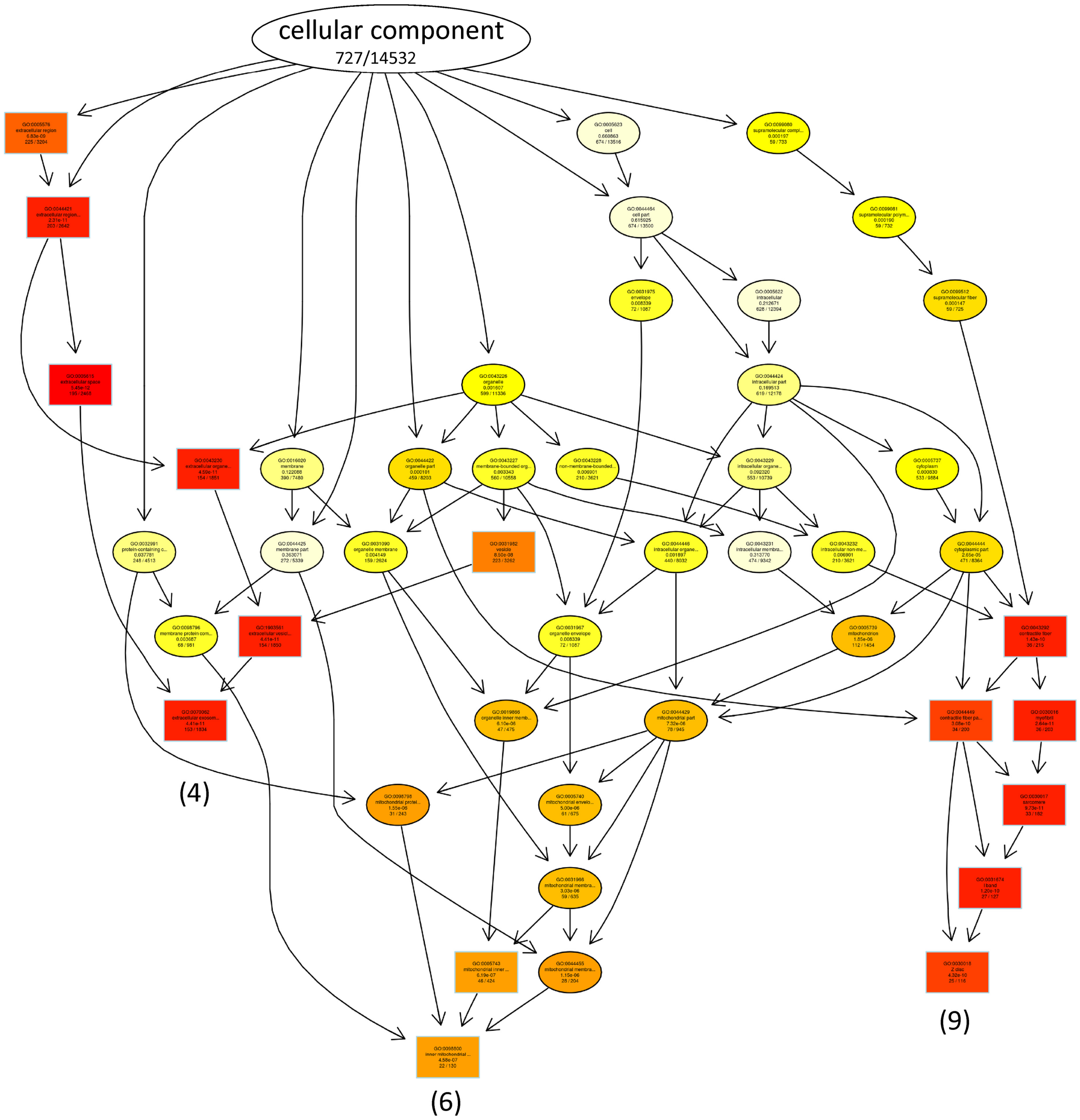


**Supplemental Figure S5: Ancestor Chart created for the 15 most significant GO terms in the category cellular component.**

The 15 most significant GO terms were displayed as squares, the connected GO terms as circles. The color is defined by the significance illustrating the most significant terms in red.

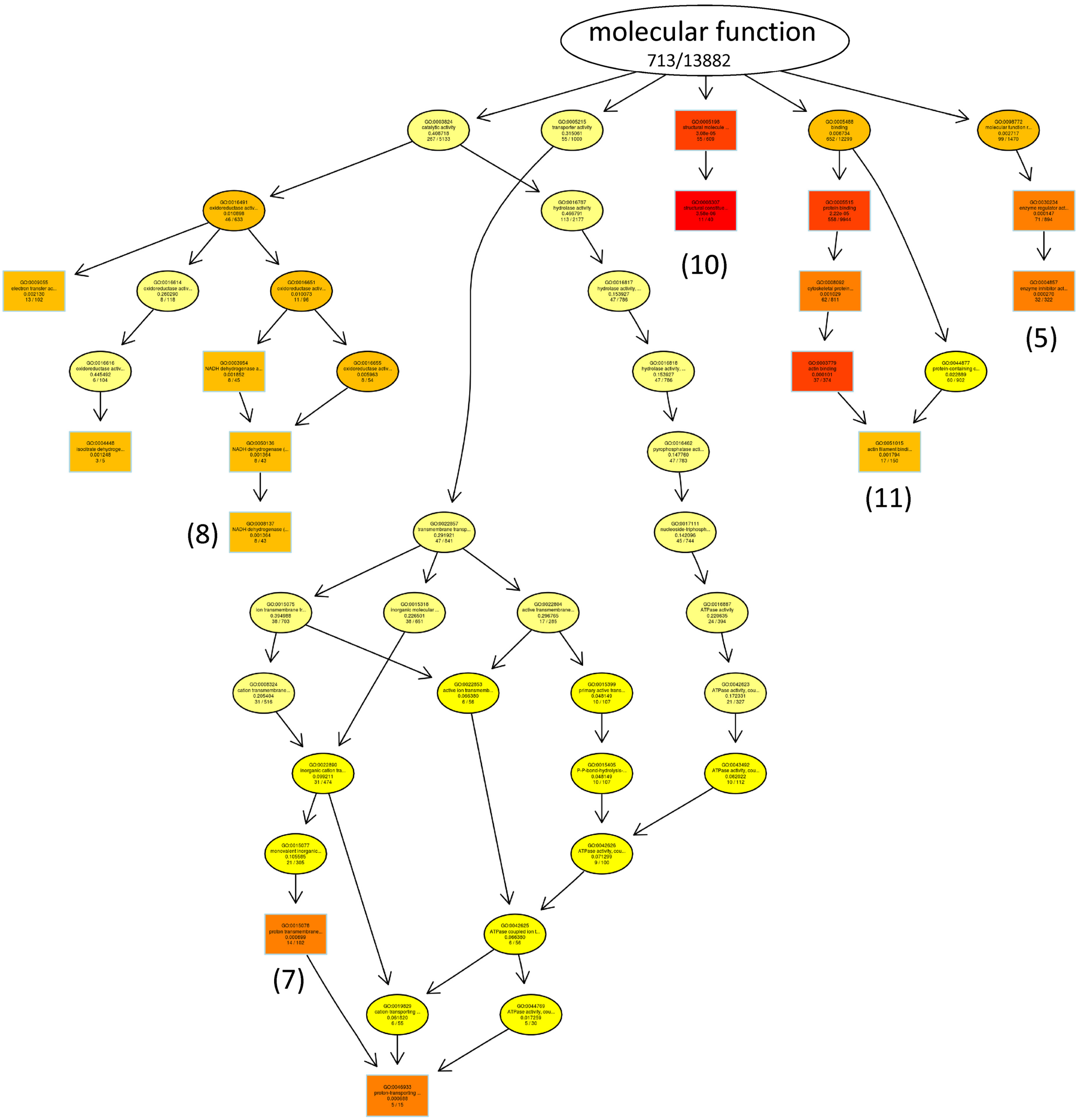


**Supplemental Figure S6: Ancestor Chart created for the 15 most significant GO terms in the category molecular function.**

The 15 most significant GO terms were displayed as squares, the connected GO terms as circles. The color is defined by the significance illustrating the most significant terms in red. For number (7) the parental GO term was selected because in the most significant GO term only 15 genes were annotated.

**Supplemental Table S5: List of significantly regulated genes displayed in the GO circle in Figure 5E.**

| GO term information | Significantly upregulated genes | Significantly downregulated genes |
|---|---|---|
| **defense response to virus**<br>GO:0051607<br>Annotated genes: 190<br>Significant regulated genes: 29<br>p-value: $1.0 \cdot 10^{-7}$ | *ADAR - BIRC3 - BST2 - DTX3L - EIF2AK2 - GBP1 - IFI44L - IFIT1 - IFIT3 - IRF1 - IRF2 - IRF7 - ISG15 - MUL1 - MX1 - MX2 - OAS3 - PARP9 - PLSCR1 - RNF26 - RSAD2 - STAT1 - TBK1 - TNFAIP3 - TRIM25 - TRIM56* | *BNIP3 - IL33 - POLR3D* |
| **immune system process**<br>GO:0002376<br>Annotated genes: 2376<br>Significant regulated genes: 170<br>p-value: $7.6 \cdot 10^{-7}$ | *ADAR - ADD2 - ADGRG3 - AGER - ALDH3B1 - ANXA1 - APOD - APOL1 - AQP4 - ATAD3B - AZI2 - B2M - BCL6 - BIRC3 - BPIFB1 - BRAF - BRCA2 - BST2 - BTN3A2 - C1RL - C3 - CASP4 - CCL20 - CD177 - CD34 - CD48 - CFB - CFD - CLU - CMTM6 - CREBBP - CSF3 - CSK - CTSH - CXCL1 - CXCL16 - CXCL2 - DCD - DDX3X - DTX3L - EIF2AK2 - ELF2 - EMP2 - EPCAM - GBP1 - GBP4 - GSDMD - HERC6 - HLA-B - HLA-C - HLA-DMA - HLA-DPB1 - HLA-E - HP - HSPA1A - HSPA6 - HSPH1 - ICAM1 - IDH1 - IFI35 - IFI44L - IFI6 - IFIT1 - IFIT3 - IL1R2 - IL6R - INPP5D - IRF1 - IRF2 - IRF7 - ISG15 - LAT2 - LCN2 - LRG1 - LTB - LTBR - MUC1 - MUC5B - MUL1 - MX1 - MX2 - N4BP2L2 - OAS3 - PARP9 - PILRB - PLSCR1 - POU2F2 - PPBP - PSMB9 - RAB34 - RB1 - RHOF - RNF26 - RSAD2 - RUBCN - SBSPON - SECTM1 - SLC2A3 - SLC8B1 - SLPI - SPINK5 - SPN - STAT1 - TAP1 - TBK1 - TF - TMEM190 - TMEM91 - TNFAIP3 - TNFAIP6 - TNFRSF10A - TNFRSF14 - TNFRSF1B - TNFSF10 - TNFSF14 - TRIM25 - TRIM56 - TYROBP - VAMP3 - VEGFD - XAF1* | *ABI1 - ACTN1 - ADA2 - ADGRE5 - ALDOA - ANO6 - AP1S2 - ARID5A - ARPC1A - ATP1B1 - BNIP3 - C6 - CACNA1C - CD63 - CDC73 - CDKN1A - CMKLR1 - COL1A1 - COL1A2 - CYFIP2 - DAPK2 - DSP - EPRS - EPS8 - ERAP1 - EXOSC9 - FKBP1A - FN1 - FUCA2 - GAPDH - HK1 - HSPA9 - IL33 - INPPL1 - LTF - LYZ - MB - NPPA - POLR3D - PPP1R14B - PRDX6 - PRKRA - RAB8B - SEC23A - SLC16A1 - SNX4 - TESC - VCL - ZNF160* |
| **innate immune response**<br>GO:0045087<br>Annotated genes: 710<br>Significant regulated genes: 66<br>p-value: $1.2 \cdot 10^{-6}$ | *ADAR - AGER - ANXA1 - APOL1 - AQP4 - B2M - BIRC3 - BPIFB1 - BST2 - C1RL - C3 - CASP4 - CCL20 - CD177 - CFB - CFD - CLU - CREBBP - CSK - CXCL16 - DDX3X - DTX3L - EIF2AK2 - GBP1 - GBP4 - GSDMD - HLA-B - HLA-C - HLA-DPB1 - HSPA1A - ICAM1 - IFI35 - IFI6 - IFIT1 - IFIT3 - IRF1 - IRF2 - IRF7 - ISG15 - LCN2 - MUC1 - MUC5B - MUL1 - MX1 - MX2 - OAS3 - PARP9 - PLSCR1 - RSAD2 - SLPI - STAT1 - TBK1 - TNFAIP3 - TRIM25 - TRIM56 - TYROBP - VAMP3 - XAF1* | *ARID5A - C6 - EPRS - ERAP1 - GAPDH - LTF - POLR3D - PPP1R14B* |
| **antimicrobial humoral response**<br>GO:0019730<br>Annotated genes: 71<br>Significant regulated genes: 9<br>p-value: 0.00951 | *BPIFB1 - CLU - DCD - LCN2 - SLPI - SPINK5* | *GAPDH - LTF - LYZ* |

| | | |
|---|---|---|
| **positive regulation of I-kappaB kinase/NF-kappaB signaling**<br>GO:0043123<br>Annotated genes: 154<br>Significant regulated genes: 13<br>p-value: 0.04993 | *BIRC3 - BST2 - CARD16 - CFLAR - LTBR - MUL1 - SECTM1 - SHISA5 - TBK1 - TNFSF10 - TRIM25* | *FKBP1A - LTF* |
| **extracellular space**<br>GO:0005615<br>Annotated genes: 2468<br>Significant regulated genes: 195<br>p-value: $5.5 \cdot 10^{-12}$ | *ADIRF - AHCTF1 - ALDH3B1 - ANXA1 - APOC1 - APOD - APOE - APOL1 - APPL2 - B2M - BPIFB1 - BST2 - C1RL - C3 - CA4 - CCL20 - CD177 - CD48 - CD82 - CFB - CFD - CLU - COBLL1 - CPNE8 - CRNN - CSF3 - CSK - CTSH - CXCL1 - CXCL16 - CXCL2 - DCD - DDX3X - DLK1 - DNAJA1 - DNAJB1 - EPCAM - EPHB4 - FGF18 - FRZB - GPIHBP1 - GPM6A - GSDMD - HIST1H2AC - HLA-B - HLA-C - HP - HPR - HSPA1A - HSPA6 - HSPH1 - ICAM1 - IDH1 - IGFBP4 - JADE2 - KRT19 - LAP3 - LAT2 - LCN2 - LGI4 - LRG1 - LRRC57 - LTB - MAL2 - MAN1A1 - MEST - MSMB - MUC1 - MUC5B - N4BP2L2 - NAPSA - OAS3 - PARP4 - PLSCR1 - PPBP - PRSS8 - PSCA - PSMB9 - PTPRD - RAB34 - RAPGEF3 - RHOF - SCGB3A1 - SECTM1 - SFN - SFTPB - SLC2A3 - SLPI - SORL1 - SPN - SRI - STX2 - TF - TM7SF3 - TNFAIP3 - TNFAIP6 - TNFSF10 - TNFSF14 - TPM3 - VASH1 - VEGFD - WARS - WFDC2* | *ABI1 - ACTN1 - ACTN2 - ADA2 - ADGRE5 - ALDOA - ANO6 - ARPC1A - ATP1B1 - ATP5F1A - ATP5F1B - AZGP1 - BGN - C6 - CD63 - CDH13 - CKB - COL1A1 - COL1A2 - CRYAB - CS - CSE1L - CYB5R1 - CYFIP2 - DSP - DSTN - ENO3 - EPS8 - ERAP1 - EXOSC9 - F13A1 - F5 - FN1 - FUCA2 - GAPDH - GARS - GOT2 - HJV - HNRNPA1 - HSPA9 - IDH2 - IL33 - KALRN - LACRT - LAMC1 - LCN1 - LOXL1 - LTF - LYZ - MB - MGP - NAPG - NCL - NEBL - NPPA - NPPB - OPRPN - PACSIN3 - PDCD5 - PGK1 - PIP - POSTN - PRDX6 - RAB21 - RAB8B - RHOB - RHOQ - RPL12 - RPLP2 - RPS8 - RSU1 - SCGB1D1 - SCGB2A1 - SLC16A1 - SLC25A3 - SMPD1 - SMS - SND1 - SOD3 - THNSL2 - TIMM8B - TIMP1 - TMEM256 - TNC - TTN - UBE2D2 - UBE2G1 - VCL - VDAC1 - VPS4A - WWP1 - YWHAG* |
| **extracellular exosome**<br>GO:0070062<br>Annotated genes: 1834<br>Significant regulated genes: 153<br>p-value: $4.4 \cdot 10^{-11}$ | *ADIRF - AHCTF1 - ALDH3B1 - ANXA1 - APOD - APOE - APPL2 - B2M - BPIFB1 - BST2 - C1RL - C3 - CA4 - CD177 - CD48 - CD82 - CFB - CFD - CLU - COBLL1 - CPNE8 - CRNN - CSK - CTSH - DCD - DDX3X - DNAJA1 - DNAJB1 - EPCAM - EPHB4 - GPM6A - HIST1H2AC - HLA-B - HLA-C - HP - HPR - HSPA1A - HSPA6 - HSPH1 - ICAM1 - IDH1 - JADE2 - KRT19 - LAP3 - LAT2 - LCN2 - LRG1 - LRRC57 - MAL2 - MAN1A1 - MEST - MUC1 - MUC5B - N4BP2L2 - NAPSA - PARP4 - PLSCR1 - PRSS8 - PSCA - PSMB9 - PTPRD - RAB34 - RAPGEF3 - RHOF - SCGB3A1 - SECTM1 - SFN - SLC2A3 - SLPI - SORL1 - SPN - SRI - TF - TM7SF3 - TNFAIP3 - TNFSF10 - TPM3 - WARS - WFDC2* | *ABI1 - ACTN1 - ACTN2 - ADGRE5 - ALDOA - ANO6 - ARPC1A - ATP1B1 - ATP5F1A - ATP5F1B - AZGP1 - BGN - C6 - CD63 - CDH13 - CKB - COL1A2 - CRYAB - CS - CSE1L - CYB5R1 - CYFIP2 - DSP - DSTN - ENO3 - EPS8 - ERAP1 - EXOSC9 - FN1 - FUCA2 - GAPDH - GARS - GOT2 - HNRNPA1 - HSPA9 - IDH2 - KALRN - LAMC1 - LTF - LYZ - MB - MGP - NAPG - NCL - NEBL - PACSIN3 - PDCD5 - PGK1 - PIP - PRDX6 - RAB21 - RAB8B - RHOB - RHOQ - RPL12 - RPLP2 - RPS8 - RSU1 - SLC16A1 - SLC25A3 - SMPD1 - SMS - SND1 - SOD3 - TIMP1 - TMEM256 - TTN - UBE2D2 - UBE2G1 - VCL - VDAC1 - VPS4A - WWP1 - YWHAG* |
| **collagen-containing extracellular matrix**<br>GO:0062023<br>Annotated genes: 219<br>Significant regulated genes: 23<br>p-value: 0.00063 | *APOE - CLU - COL27A1 - ICAM1 - LAMB3 - SBSPON - SLPI* | *ASPN - AZGP1 - BGN - CCDC80 - CDH13 - COL1A1 - COL1A2 - FN1 - LAMC1 - LOXL1 - MGP - NPPA - POSTN - SOD3 - TIMP1 - TNC* |
| **blood microparticle**<br>GO:0072562<br>Annotated genes: 100 | *APOE - APOL1 - C3 - CFB - CLU - HP - HPR - HSPA1A - HSPA6 - TF* | *F13A1 - FN1* |

| | | |
|---|---|---|
| Significant regulated genes: 12<br>p-value: 0.00415 | | |
| **mitochondrion**<br>GO:0005739<br>Annotated genes: 1454<br>Significant regulated genes: 112<br>p-value: $1.8 \cdot 10^{-6}$ | *ABCA8 - ABCG1 - ANXA1 - ATAD3B - ATP5PD - BAG5 - BRAF - CASP4 - CLU - DDX3X - DNAJA1 - G0S2 - HIF3A - HSPA1A - IDH1 - IFI6 - IFIT3 - MIGA2 - MIPEP - MTERF2 - MUL1 - MX1 - MX2 - MXD1 - PARP9 - RSAD2 - SCO2 - SFN - SLC25A28 - SLC8B1 - SRI - TK2 - TYMP - UBIAD1 - WDR81 - XAF1* | *AFG3L2 - APEX1 - APEX2 - ATP5F1A - ATP5F1B - ATP5MC3 - ATP5MG - ATP5MPL - ATP5PF - ATPAF1 - BCKDK - BNIP3 - BOLA3 - CARD19 - CHCHD10 - CHPF - CKMT2 - COX4I1 - COX5B - COX6A2 - COX6B1 - COX6C - CREB3L4 - CRYAB - CS - CYB5R1 - DLD - DNM1L - ECHS1 - ETFA - ETFB - FAM162A - FECH - GARS - GATD3A - GOT2 - GSK3B - HADH - HADHB - HK1 - HSPA9 - IDH2 - IDH3G - LDHD - MLYCD - MRPL13 - MRPL15 - MRPL32 - MRPS12 - MRPS17 - MRPS33 - MSRB2 - NAPG - NDRG4 - NDUFA10 - NDUFA9 - NDUFB1 - NDUFB8 - NDUFS1 - NDUFS3 - NDUFV2 - NDUFV3 - NOC3L - PLIN5 - PRELID3B - SLC25A12 - SLC25A3 - SLC25A4 - TCHP - TIMM17A - TIMM8B - TRIT1 - UQCRFS1 - UQCRH - VDAC1 - YWHAG* |
| **identical protein binding**<br>GO:0042802<br>Annotated genes: 1476<br>Significant regulated genes: 96<br>p-value: 0.00842 | *ABCG1 - ADD2 - AGER - ANXA1 - APOE - ARL6IP1 - B2M - BCL6 - BRAF - BRCA2 - BST2 - CARD16 - CLDN15 - CLDN4 - COIL - CRLF3 - CSK - EIF2AK2 - GBP1 - GBP4 - ID1 - IDH1 - IFIT3 - IL6R - LTBR - MIGA2 - MMACHC - MUL1 - MVD - MX1 - MZF1 - RB1 - SFN - SLC8B1 - SRGAP2C - STAT1 - TAP1 - TBK1 - TMEM192 - TNFAIP3 - TNFSF10 - TYROBP - TYRP1 - VEGFD - WARS* | *ACTN1 - ACTN2 - ADA2 - ALDOA - ANO6 - APCDD1 - BNIP3 - CAP2 - CBY1 - CDH13 - COL1A1 - COL1A2 - CRYAB - CSRP3 - DAPK2 - DNM1L - ENO3 - EPRS - ESYT2 - FHL2 - FKBP1A - FN1 - GAPDH - GARS - GNPNAT1 - GOT2 - HK1 - HSPB6 - HSPB8 - KCNK1 - LYZ - MLYCD - MTUS2 - NCL - PLIN5 - PRDX6 - PRKRA - PSME3 - RPL7 - SLC16A1 - SLC25A12 - TENM2 - TERF2 - TESC - TNNC1 - TPM1 - TPM2 - TTN - USP2 - VDAC1 - YWHAG* |
| **regulation of apoptotic process**<br>GO:0042981<br>Annotated genes: 1314<br>Significant regulated genes: 84<br>p-value: 0.0155 | *ADAR - ANXA1 - APOE - ARHGEF1 - ARL6IP1 - BAG5 - BCL6 - BIRC3 - BRAF - CARD16 - CASP4 - CFLAR - CLU - CREBBP - CTSH - DDX3X - DEDD2 - DHCR24 - DNAJA1 - EIF2AK2 - FRZB - G0S2 - GRINA - HSPA1A - HSPH1 - ICAM1 - ID1 - IFI6 - IFIT3 - INPP5D - IRF7 - KCNB1 - LTBR - MUC1 - MUL1 - NUPR1 - SFN - STAT1 - TNFAIP3 - TNFRSF10A - TNFRSF14 - TNFRSF1B - TNFSF10 - TNFSF14* | *AAMDC - ACTN1 - ACTN2 - ANKRD1 - ANO6 - APEX1 - BNIP3 - CDC73 - CDKN1A - CRYAB - DAB2 - DAPK2 - DNM1L - EGLN3 - FAM162A - FHL2 - GAPDH - GSK3B - HSPA9 - HSPB6 - ITSN1 - KALRN - LACRT - LTF - MLLT11 - NDUFS3 - PDCD5 - PDE3A - PIP - PRKRA - PSME3 - QRICH1 - RETREG1 - RHOB - RRN3 - SH3RF2 - SMPD1 - SRSF6 - TIMP1 - YWHAG* |

**Supplemental Table S6: List of significantly regulated genes displayed in the GO circle in Figure 6A.**

| GO term information | Significantly upregulated genes | Significantly downregulated genes |
|---|---|---|
| **type I interferon signaling pathway**<br>GO:0060337<br>Annotated genes: 69<br>Significant regulated genes: 19<br>p-value: $8.5 \cdot 10^{-10}$ | *ADAR - BST2 - HLA-B - HLA-C - IFI35 - IFI6 - IFIT1 - IFIT3 - IRF1 - IRF2 - IRF7 - ISG15 - MUL1 - MX1 - MX2 - OAS3 - RSAD2 - STAT1 - XAF1* | - |
| **defense response to virus**<br>GO:0051607<br>Annotated genes: 190<br>Significant regulated genes: 29<br>p-value: $1.0 \cdot 10^{-7}$ | *ADAR - BIRC3 - BST2 - DTX3L - EIF2AK2 - GBP1 - IFI44L - IFIT1 - IFIT3 - IRF1 - IRF2 - IRF7 - ISG15 - MUL1 - MX1 - MX2 - OAS3 - PARP9 - PLSCR1 - RNF26 - RSAD2 - STAT1 - TBK1 - TNFAIP3 - TRIM25 - TRIM56* | *BNIP3 - IL33 - POLR3D* |
| **negative regulation of viral genome replication**<br>GO:0045071<br>Annotated genes: 47<br>Significant regulated genes: 13<br>p-value: $3.8 \cdot 10^{-7}$ | *ADAR - BST2 - EIF2AK2 - IFIT1 - ISG15 - MX1 - OAS3 - PARP10 - PLSCR1 - PROX1 - RSAD2 - SLPI* | *LTF* |
| **extracellular exosome**<br>GO:0070062<br>Annotated genes: 1834<br>Significant regulated genes: 153<br>p-value: $4.4 \cdot 10^{-11}$ | *ADIRF - AHCTF1 - ALDH3B1 - ANXA1 - APOD - APOE - APPL2 - B2M - BPIFB1 - BST2 - C1RL - C3 - CA4 - CD177 - CD48 - CD82 - CFB - CFD - CLU - COBLL1 - CPNE8 - CRNN - CSK - CTSH - DCD - DDX3X - DNAJA1 - DNAJB1 - EPCAM - EPHB4 - GPM6A - HIST1H2AC - HLA-B - HLA-C - HP - HPR - HSPA1A - HSPA6 - HSPH1 - ICAM1 - IDH1 - JADE2 - KRT19 - LAP3 - LAT2 - LCN2 - LRG1 - LRRC57 - MAL2 - MAN1A1 - MEST - MUC1 - MUC5B - N4BP2L2 - NAPSA - PARP4 - PLSCR1 - PRSS8 - PSCA - PSMB9 - PTPRD - RAB34 - RAPGEF3 - RHOF - SCGB3A1 - SECTM1 - SFN - SLC2A3 - SLPI - SORL1 - SPN - SRI - TF - TM7SF3 - TNFAIP3 - TNFSF10 - TPM3 - WARS - WFDC2* | *ABI1 - ACTN1 - ACTN2 - ADGRE5 - ALDOA - ANO6 - ARPC1A - ATP1B1 - ATP5F1A - ATP5F1B - AZGP1 - BGN - C6 - CD63 - CDH13 - CKB - COL1A2 - CRYAB - CS - CSE1L - CYB5R1 - CYFIP2 - DSP - DSTN - ENO3 - EPS8 - ERAP1 - EXOSC9 - FN1 - FUCA2 - GAPDH - GARS - GOT2 - HNRNPA1 - HSPA9 - IDH2 - KALRN - LAMC1 - LTF - LYZ - MB - MGP - NAPG - NCL - NEBL - PACSIN3 - PDCD5 - PGK1 - PIP - PRDX6 - RAB21 - RAB8B - RHOB - RHOQ - RPL12 - RPLP2 - RPS8 - RSU1 - SLC16A1 - SLC25A3 - SMPD1 - SMS - SND1 - SOD3 - TIMP1 - TMEM256 - TTN - UBE2D2 - UBE2G1 - VCL - VDAC1 - VPS4A - WWP1 - YWHAG* |
| **enzyme inhibitor activity**<br>GO:0004857<br>Annotated genes: 322<br>Significant regulated genes: 32<br>p-value: 0.00027 | *ANXA1 - APOC1 - BIRC3 - BST2 - C3 - CARD16 - DTX3L - PARP9 - PDE6G - SAG - SFN - SLPI - SPINK5 - TF - TNFSF14 - WARS - WFDC2* | *ASPN - BGN - CDKN1A - GAPDH - LCN1 - LTF - OPRPN - PKIA - PKIG - PPP1R14B - SH3RF2 - SLN - TESC - TIMP1 - YWHAG* |
| **inner mitochondrial membrane protein complex**<br>GO:0098800<br>Annotated genes: 130 | *ATP5PD* | *AFG3L2 - ATP5F1A - ATP5F1B - ATP5MC3 - ATP5MG - ATP5MPL - ATP5PF - COX4I1 - COX5B - COX6A2 - NDUFA10 - NDUFA9 - NDUFB1 -* |

| | | |
|---|---|---|
| Significant regulated genes: 22<br>p-value: $4.6 \cdot 10^{-7}$ | | *NDUFB8 - NDUFS1 - NDUFS3 - NDUFV2 - NDUFV3 - TIMM17A - UQCRFS1 - UQCRH* |
| **proton transmembrane transporter activity**<br>GO:0015078<br>Annotated genes: 102<br>Significant regulated genes: 14<br>p-value: 0.0007 | *ATP5PD - SLC17A5* | *ATP5F1A - ATP5F1B - ATP5MC3 - ATP5MG - ATP5PF - COX4I1 - COX5B - COX6A2 - COX6B1 - COX6C - SLC25A3 - SLC9A8* |
| **NADH dehydrogenase (ubiquinone) activity**<br>GO:0008137<br>Annotated genes: 43<br>Significant regulated genes: 8<br>p-value: 0.00136 | - | *NDUFA10 - NDUFA9 - NDUFB1 - NDUFB8 - NDUFS1 - NDUFS3 - NDUFV2 - NDUFV3* |
| **Z disc**<br>GO:0030018<br>Annotated genes: 146<br>Significant regulated genes: 25<br>p-value: $4.3 \cdot 10^{-10}$ | *KRT19 - MYBPC1 - SRI* | *ACTN1 - ACTN2 - CACNA1C - CRYAB - CSRP3 - FBXO32 - FHL2 - FHOD3 - FKBP1A - FLNC - HRC - ITGB1BP2 - MYH7 - MYPN - MYZAP - NEBL - NEXN - PARVB - PDLIM3 - PPP1R12B - TCAP - TTN* |
| **structural constituent of muscle**<br>GO:0008307<br>Annotated genes: 40<br>Significant regulated genes: 11<br>p-value: $3.6 \cdot 10^{-6}$ | *KRT19 - MYBPC1 - TPM3* | *ACTN2 - CSRP3 - NEBL - NEXN - TCAP - TPM1 - TPM2 - TTN* |
| **actin filament binding**<br>GO:0051015<br>Annotated genes: 150<br>Significant regulated genes: 17<br>p-value: 0.00179 | *ADD2 - MYBPC1 - TPM3* | *ACTN1 - ACTN2 - ARPC1A - DSTN - FLNC - MYH7 - MYPN - NEBL - NEXN - TNNC1 - TPM1 - TPM2 - TTN - VCL* |

**Supplemental Table S7: List of significantly regulated genes displayed in the GO circle in Figure 6E: response to virus.**

| GO term information | Significantly upregulated genes | Significantly downregulated genes |
|---|---|---|
| **defense response to virus**<br>GO:0051607<br>Annotated genes: 190<br>Significant regulated genes: 39<br>p-value: $1.0 \cdot 10^{-7}$ | *ADAR - BIRC3 - BST2 - DTX3L - EIF2AK2 - GBP1 - IFI44L - IFIT1 - IFIT3 - IRF1 - IRF2 - IRF7 - ISG15 - MUL1 - MX1 - MX2 - OAS3 - PARP9 - PLSCR1 - RNF26 - RSAD2 - STAT1 - TBK1 - TNFAIP3 - TRIM25 - TRIM56* | *BNIP3 - IL33 - POLR3D* |
| **response to virus**<br>GO:0009615<br>Annotated genes: 265<br>Significant regulated genes: 35<br>p-value: $2.0 \cdot 10^{-7}$ | *ADAR - BIRC3 - BST2 - CLU - DDX3X - DTX3L - EIF2AK2 - GBP1 - IFI44 - IFI44L - IFIT1 - IFIT3 - IRF1 - IRF2 - IRF7 - ISG15 - MUL1 - MX1 - MX2 - OAS3 - PARP9 - PLSCR1 - RNF26 - RSAD2 - STAT1 - TBK1 - TNFAIP3 - TRIM25 - TRIM56* | *BNIP3 - DDX1 - DDX21 - IL33 - POLR3D - PRKRA* |
| **negative regulation of viral genome replication**<br>GO:0045071<br>Annotated genes: 47<br>Significant regulated genes: 13<br>p-value: $3.8 \cdot 10^{-7}$ | *ADAR - BST2 - EIF2AK2 - IFIT1 - ISG15 - MX1 - OAS3 - PARP10 - PLSCR1 - PROX1 - RSAD2 - SLPI* | *LTF* |
| **negative regulation of viral life cycle**<br>GO:1903901<br>Annotated genes: 65<br>Significant regulated genes: 14<br>p-value: $3.7 \cdot 10^{-6}$ | *ADAR - BST2 - EIF2AK2 - IFIT1 - ISG15 - MX1 - OAS3 - PARP10 - PLSCR1 - PROX1 - RSAD2 - SLPI - TRIM25* | *LTF* |
| **negative regulation of viral process**<br>GO:0048525<br>Annotated genes: 76<br>Significant regulated genes: 15<br>p-value: $5.2 \cdot 10^{-6}$ | *ADAR - BST2 - EIF2AK2 - IFIT1 - ISG15 - MX1 - OAS3 - PARP10 - PLSCR1 - PROX1 - RSAD2 - SLPI - STAT1 - TRIM25* | *LTF* |
| **regulation of viral genome replication**<br>GO:0045069<br>Annotated genes: 79<br>Significant regulated genes: 14<br>p-value: $3.9 \cdot 10^{-5}$ | *ADAR - BST2 - DDX3X - EIF2AK2 - IFIT1 - ISG15 - MX1 - OAS3 - PARP10 - PLSCR1 - PROX1 - RSAD2 - SLPI* | *LTF* |
| **regulation of viral process**<br>GO:0050792<br>Annotated genes: 154 | *ADAR - APOE - BST2 - DDX3X - EIF2AK2 - GTF2B - IFIT1 - ISG15 - MX1 - OAS3 - PARP10 - PLSCR1 - PROX1 - RSAD2 - SLPI - STAT1 - TRIM25* | *LTF - VPS4A* |

| | | |
|---|---|---|
| Significant regulated genes: 19<br>p-value: 0.0003 | | |
| **regulation of viral life cycle**<br>GO:1903900<br>Annotated genes: 119<br>Significant regulated genes: 16<br>p-value: 0.00033 | *ADAR - BST2 - DDX3X - EIF2AK2 - IFIT1 - ISG15 - MX1 - OAS3 - PARP10 - PLSCR1 - PROX1 - RSAD2 - SLPI - TRIM25* | *LTF - VPS4A* |
| **viral process**<br>GO:0016032<br>Annotated genes: 633<br>Significant regulated genes: 52<br>p-value: 0.00041 | *ADAR - APOE - AZI2 - B2M - BST2 - CFLAR - CREBBP - CUL5 - DDX3X - E4F1 - EIF2AK2 - GTF2B - HLA-B - HLA-C - HSPA1A - ICAM1 - IFIT1 - IRF7 - ISG15 - KRT19 - LTBR - MX1 - OAS3 - PARP10 - PLSCR1 - PROX1 - PSMB9 - RB1 - RSAD2 - SLPI - STAT1 - TAF1 - TAP1 - TBK1 - TNFRSF14 - TRIM25* | *ABI1 - AP1S2 - BNIP3 - CPSF4 - EIF3G - ELOC - HNRNPA1 - ITSN1 - KPNA4 - LTF - MORC3 - SLC25A4 - SND1 - VDAC1 - VPS4A - WWP1* |
| **viral genome replication**<br>GO:0019079<br>Annotated genes: 105<br>Significant regulated genes: 14<br>p-value: 0.00084 | *ADAR - BST2 - DDX3X - EIF2AK2 - IFIT1 - ISG15 - MX1 - OAS3 - PARP10 - PLSCR1 - PROX1 - RSAD2 - SLPI* | *LTF* |
| **regulation of defense response to virus by host**<br>GO:0050691<br>Annotated genes: 33<br>Significant regulated genes: 6<br>p-value: 0.00575 | *DTX3L - IFIT1 - MUL1 - PARP9 - STAT1 - TNFAIP3* | - |
| **regulation of defense response to virus**<br>GO:0050688<br>Annotated genes: 64<br>Significant regulated genes: 8<br>p-value: 0.01528 | *BIRC3 - DTX3L - IFIT1 - MUL1 - PARP9 - RNF26 - STAT1 - TNFAIP3* | - |
| **viral life cycle**<br>GO:0019058<br>Annotated genes: 259<br>Significant regulated genes: 21<br>p-value: 0.02344 | *ADAR - APOE - BST2 - DDX3X - EIF2AK2 - HSPA1A - ICAM1 - IFIT1 - ISG15 - MX1 - OAS3 - PARP10 - PLSCR1 - PROX1 - RSAD2 - SLPI - TNFRSF14 - TRIM25* | *LTF - VPS4A - WWP1* |

**Supplemental Table S8: List of significantly regulated genes displayed in the GO circle in Figure 6F: on the right side platelet & complement / on the left side lipoproteins.**

| GO term information | Significantly upregulated genes | Significantly downregulated genes |
|---|---|---|
| **platelet alpha granule lumen**<br>GO:0031093<br>Annotated genes: 63<br>Significant regulated genes: 11<br>p-value: 0.00026 | *CFD - CLU - PPBP - VEGFD* | *ACTN1 - ACTN2 - ALDOA - F13A1 - F5 - FN1 - TIMP1* |
| **platelet degranulation**<br>GO:0002576<br>Annotated genes: 118<br>Significant regulated genes: 16<br>p-value: 0.0003 | *CFD - CLU - PPBP - TF - VEGFD* | *ACTN1 - ACTN2 - ALDOA - CD63 - CYB5R1 - F13A1 - F5 - FN1 - TIMP1 - TTN - VCL* |
| **platelet alpha granule**<br>GO:0031091<br>Annotated genes: 86<br>Significant regulated genes: 11<br>p-value: 0.00026 | *CFD - CLU - PPBP - VEGFD* | *ACTN1 - ACTN2 - ALDOA - CYB5R1 - F13A1 - F5 - FN1 - TIMP1* |
| **complement activation, alternative pathway**<br>GO:0006957<br>Annotated genes: 13<br>Significant regulated genes: 3<br>p-value: 0.02548 | *C3 - CFB - CFD* | - |
| **high-density lipoprotein particle**<br>GO:0034364<br>Annotated genes: 21<br>Significant regulated genes: 6<br>p-value: 0.00044 | *APOC1 - APOE - APOL1 - CLU - GPIHBP1 - HPR* | - |
| **lipoprotein particle / plasma lipoprotein particle**<br>GO:1990777 / GO:0034358<br>Annotated genes: 33<br>Significant regulated genes: 7<br>p-value: 0.00104 | *APOC1 - APOE - APOL1 - CLU - GPIHBP1 - HPR - SORL1* | - |
| **lipoprotein particle binding**<br>GO:0071813<br>Annotated genes: 22 | *APOE - GPIHBP1 - SORL1* | *CDH13* |

| | | |
|---|---|---|
| Significant regulated genes: 4<br>p-value: 0.02411 | | |
| **high-density lipoprotein particle remodeling**<br>GO:0034375<br>Annotated genes: 14<br>Significant regulated genes: 3<br>p-value: 0.03123 | *ABCG1 - APOC1 - APOE* | - |
| **very-low-density lipoprotein particle / triglyceride-rich plasma lipoprotein particle**<br>GO:0034361 / GO:0034385<br>Annotated genes: 16<br>Significant regulated genes: 3<br>p-value: 0.0429 | *APOC1 - APOE - APOL1* | - |

**Supplemental Table S9: List of significantly regulated genes displayed in the GO circle in Figure 6G: MHC antigene presentation.**

| GO term information | Significantly upregulated genes | Significantly downregulated genes |
|---|---|---|
| **MHC protein complex**<br>GO:0042611<br>Annotated genes: 22<br>Significant regulated genes: 6<br>p-value: 0.00057 | *B2M - HLA-B - HLA-C - HLA-DMA - HLA-DPB1 - HLA-E* | - |
| **MHC protein binding**<br>GO:0042287<br>Annotated genes: 24<br>Significant regulated genes: 4<br>p-value: 0.03231 | *PILRB - TAP1* | *ATP5F1A - ATP5F1B* |
| **antigen processing and presentation of peptide antigen via MHC class I**<br>GO:0002474<br>Annotated genes: 45<br>Significant regulated genes: 8<br>p-value: 0.00173 | *B2M - HLA-B - HLA-C - HLA-E - TAP1 - VAMP3* | *ERAP1 - SEC23A* |
| **MHC class I protein binding**<br>GO:0042288<br>Annotated genes: 15<br>Significant regulated genes: 4<br>p-value: 0.00598 | *PILRB - TAP1* | *ATP5F1A - ATP5F1B* |
| **antigen processing and presentation of exogenous peptide antigen via MHC class I, TAP-dependent**<br>GO:0002479<br>Annotated genes: 24<br>Significant regulated genes: 5<br>p-value: 0.00633 | *B2M - HLA-B - HLA-C - TAP1 - VAMP3* | - |
| **antigen processing and presentation of exogenous peptide antigen via MHC class I**<br>GO:0042590<br>Annotated genes: 28<br>Significant regulated genes: 5<br>p-value: 0.01238 | *B2M - HLA-B - HLA-C - TAP1 - VAMP3* | - |
